# Mass mask-wearing notably reduces COVID-19 transmission

**DOI:** 10.1101/2021.06.16.21258817

**Authors:** Gavin Leech, Charlie Rogers-Smith, Jonas B. Sandbrink, Benedict Snodin, Robert Zinkov, Benjamin Rader, John S. Brownstein, Yarin Gal, Samir Bhatt, Mrinank Sharma, Sören Mindermann, Jan M. Brauner, Laurence Aitchison

## Abstract

Mask-wearing has been a controversial measure to control the COVID-19 pandemic. While masks are known to substantially reduce disease transmission in healthcare settings [1–3], studies in community settings report inconsistent results [4–6].

Investigating the inconsistency within epidemiological studies, we find that a commonly used proxy, government mask mandates, does not correlate with large increases in mask-wearing in our window of analysis. We thus analyse the effect of mask-wearing on transmission instead, drawing on several datasets covering 92 regions on 6 continents, including the largest survey of individual-level wearing behaviour (n=20 million) [7]. Using a hierarchical Bayesian model, we estimate the effect of both mask-wearing and mask-mandates on transmission by linking wearing levels (or mandates) to reported cases in each region, adjusting for mobility and non-pharmaceutical interventions.

We assess the robustness of our results in 123 experiments spanning 22 sensitivity analyses. Across these analyses, we find that an entire population wearing masks in public leads to a median reduction in the reproduction number *R* of 25.8%, with 95% of the medians between 22.2% and 30.9%. In our window of analysis, the median reduction in *R* associated with the wearing level observed in each region was 20.4% [2.0%, 23.3%]^1^. We do not find evidence that mandating mask-wearing reduces transmission. Our results suggest that mask-wearing is strongly affected by factors other than mandates.

We establish the effectiveness of mass mask-wearing, and highlight that wearing data, not mandate data, are necessary to infer this effect.

## INTRODUCTION

Face masks are one of the most prominent interventions against COVID-19, with very high uptake in most countries [7]. However, as of June 2021, global mask-wearing has begun to decline, even in countries with low vaccination rates (Figure 1). Given that only a minority of the global population is projected to be vaccinated in 2021 [8]−and given novel variants of concern that are highly transmissible and escape acquired immunity [9]−establishing the effectiveness of mask-wearing in community settings is critical. We now review past work on the effectiveness of mask-wearing in different settings and at different scales.

**Fig. 1.**
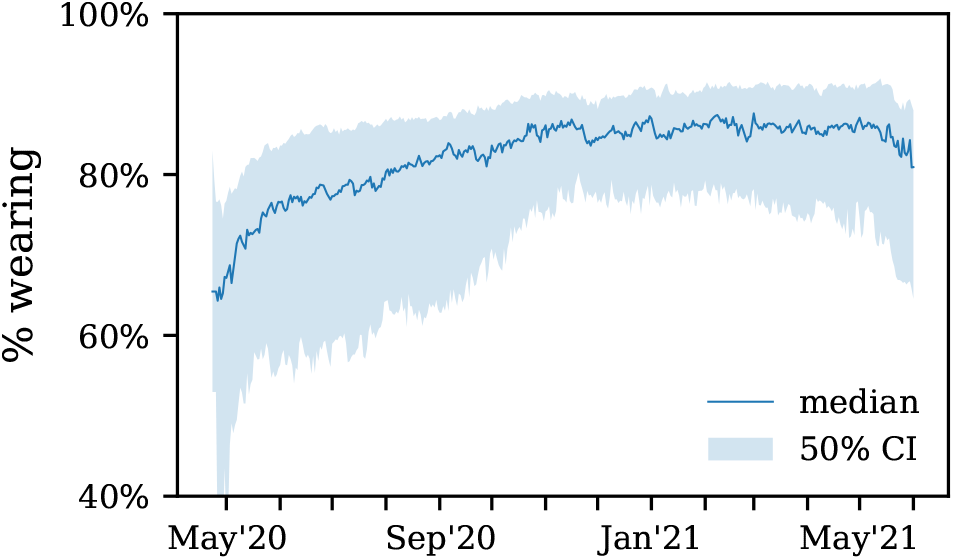
Median mask-wearing across countries in the UMD / Facebook survey [7] in which the proportion of people vaccinated as of 5th June 2021 was less than 40% (from [10]). Percentage is the proportion of people who reported that, over the last week, they wore masks most or all of the time in public spaces.

### Effects of mask-wearing in healthcare settings

In the context of healthcare, N95 masks work well when worn properly by trained users−reducing transmission of coronaviruses including SARS-CoV-2 by at least half [1, 2]. Cheng *et al*. (2021) [3] find that ideal surgical masking of a non-infected person corresponds to a 65-75% reduction in their risk of COVID-19.

### Effects of mask-wearing in small-scale community settings

Clinical studies in community settings are summarised in four meta-analyses covering SARS, COVID-19 and other respiratory infections [1, 4–6]. For fitted surgical masks, individual results from the meta-analyses range from a 7% increase in infection risk to a 61% decrease in infection risk. The meta-analytic mean decreases in infection risk vary from 4% to 15%, with large uncertainty. One of the few RCTs on mask recommendations (not mandates) found a nonsignificant and low effect [11].

Masks have at least two effects: preventing transmission to non-infected mask-wearers (‘wearer protection’), and preventing infected wearers from infecting others (‘source control’). With the exception of [3], the studies listed above estimate individual wearer protection, rather than the most policy-relevant quantity: the ecological effect of mass mask-wearing including all relevant factors. These factors include source control with average mask quality [3], the nonlinear scaling of group protection [3, 12], and risk compensation [2]. Additionally, clinical studies may not reflect the actual distribution of protection; for instance, none of the studies detailed above include cloth masks, one of the most common types [13, 14]. Finally, while mask-wearing is known to be strongly mediated by cultural factors [15–17], most studies are conducted in a specific social context and may have limited external validity.

In this study, we aim to infer the ecological effect of a large proportion of the population wearing average masks, with average fit, in the average non-residential venue, averaging across many cultures. We call this the *mass mask-wearing effect*. Our study is observational, and caution is required when making causal interpretations (see Robustness).

### Effects of mass mask-wearing, measured by mandate timing

Many studies use the timing of mask mandates as a proxy for sharp changes in the level of mass mask-wearing. Studying 41 countries, Sharma *et al*. (2020) [18] infer an inconclusive mandate effect on COVID-19 transmission centred around zero. Sharma *et al*. (2021) [19] is a regional study of 7 European countries [19] which finds an overall 7% to 17% (95% CI) reduction in transmission associated with mandates. In a mixed study of mask recommendations, mask mandates, cultural norms favouring masks, and self-reported wearing data, Leffler et al. (2020) [16] find a 26% decrease in COVID-related mortality associated with their mixed proxy for mass mask-wearing. Other studies analyse a single country: Lyu and Wehby (2020) [20] use natural experiments between US states and find a 2% absolute decrease in case growth rate after three weeks. Mitze *et al*. (2020) [21] study mandates in several regions of Germany and find a relative reduction in cases of 47%. Van Dyke *et al*. (2020) [22] exploit natural experiments between Kansas counties mandating mask-wearing and find a qualitative difference in mandating counties. In their study of US states, Chernozhukov *et al*. (2021) [23] attribute a relative ∼10% reduction in case growth rate to mandates for public-facing employees. Also studying US states, Maloney *et al*. (2020) [24] find no statistically significant change in cases following mandate implementations.

### Mandates are a poor proxy for wearing

Society-level studies of non-pharmaceutical interventions (NPIs) often use the timing of mask mandates as a proxy for wearing uptake. If mandates do not correlate with large changes in mask-wearing−for instance, due to voluntary wearing, noncompliance, or the correlation of mandate timing with (prior) support for mask-wearing−using mandate data in lieu of wearing data will lead to poor estimates of mask-wearing effectiveness.

Surprisingly, we find that national mandates may be a poor proxy for actual wearing. While Betsch *et al*. (2020) [15] find a ∼40% increase in wearing after local mandates in Germany, no other study finds a comparably large increase. In their study of US mandates, Rader *et al*. (2021) [25] did not find a statistically significant relationship between mandates and subsequent wearing. In their study of 4 US states, Adjodah *et al*. (2021) [26] find an average 23% post-mandate increase in wearing. Maloney (2020) [27] finds a 13% post-mandate increase, in US states (in the proportion ‘frequently’ or ‘always’ wearing masks). We confirm the weak correlation between mandates and subsequent wearing in 92 regions across 56 countries; see Results.

### Effects of mass mask-wearing, measured by self-reports

Instead of using mask mandates as a proxy for wearing, we use a large (n=19.97 million) global survey of mask-wearing [7]. (Our wearing covariate is also a proxy, because respondents self-report whether they have been wearing masks.) Two other studies take this approach: in their study of 24 countries, Aravindakshan *et al*. (2020) [28] use YouGov wearing data to infer an overall 3.9% to 10% relative decrease in case growth rate, for a 0-100% increase in wearing. Rader *et al*. (2021) [25] study US states using a novel SurveyMonkey wearing dataset to infer a ∼10% decrease in transmission between the lowest and highest empirical quartiles of wearing (a 50-75% increase in wearing).

Our analysis goes further than past work in the quality of wearing data−100 times the sample size, with random sampling and post-stratification−the geographical scope, the sophistication of our infection model, the incorporation of the uncertainty in epidemiological parameters, and the robustness of our results (123 sensitivity experiments).

## RESULTS

### The mandate-wearing correlation

Mask mandates are typically encoded as binary indicators that signal whether mask-wearing was required in at least some shared spaces [16, 19, 29–31]. We draw mandate data from the OxCGRT NPI database [32]. We estimate the effect of two mandate covariates and display their combined effect throughout this manuscript. The first covariate represents whether masks were ‘*required in some shared spaces, outside the home with other people present, or some situations when social distancing [was] not possible*’ (field H6 from OxCGRT, level 2 [32]). The second covariate has the same conditions, but masking is required in *all* shared spaces (field H6 from OxCGRT, level 3 or higher). Wearing estimates are from the University of Maryland / Facebook COVID-19 World Symptoms Survey [7, 33] and (for the US) the COVIDNearYou / SurveyMonkey dataset [25]. Our covariate ‘percentage of region wearing masks’ is the weighted percentage of people who said that, over the past 7 days, they wore masks in public most or all of time. The weights correct for non-response bias and for demographic imbalance [33].

Figure 2 shows the average wearing trend before and after the implementation of mandates. Most of the uptake in wearing occurs pre-mandate. In our window, the Spearman correlation coefficient between mask-wearing and mandates is *ρ* = 0.32, with *p*-value < 0.001. This is of medium strength for a correlation between social factors [34], but inadequate if mandate data is to serve as a reliable proxy of mask-wearing. This does not show that mandates do not *cause* mask-wearing, nor that there were high levels of noncompliance; it instead shows that voluntary uptake in wearing was more popular, and came earlier, than assumed in past work.

**Fig. 2.**
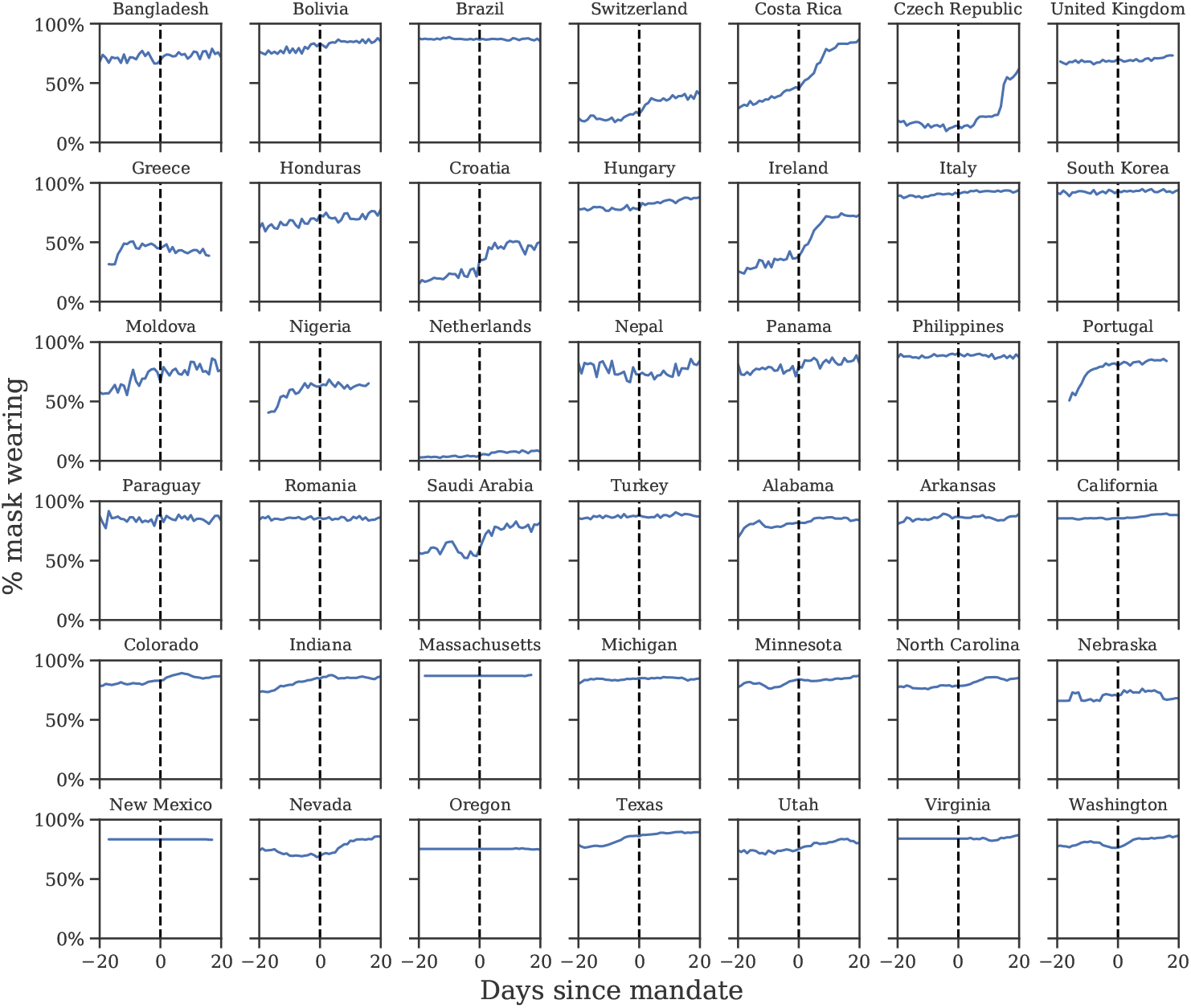
Self-reported mask-wearing against mandate timing in all regions with a new national mask mandate, May-Sep 2020. Dashed line is the date each mandate began being enforced.

Our sources of wearing data begin after April 2020−that is, after the initial transition to mask-wearing in some countries. Since it is possible that earlier mandates had persistent effects on wearing, we investigate the correlation during the first wave using an earlier YouGov wearing survey (see Appendix A). In regions with available data, most of the increase in mask-wearing occurred before the earliest national government mandates, with 64% average wearing on the day the mandate was enacted and 75% three weeks following the mandate. However, assessing the true correlation with the available data is difficult−see Discussion for details.

### Mandate and wearing effects on transmission

Using data from May to September 2020, we separately estimate the effects of mask mandates and mask-wearing in 92 regions (Table 5) with a state-of-the-art Bayesian hierarchical model (Figure 5). The model links wearing levels (or mandates) to the number of reported cases in each region via the instantaneous reproduction number *R*_*t*_. Our model is similar to [29], but in addition to adjusting for other NPIs, we also account for changes in mobility. We model many sources of uncertainty through prior distributions: epidemiological properties of the virus, differences in transmission between countries, the lag between an infection and the registration of a COVID-19 case, and the effect of unobserved influences on *R*. To obtain wearing and mandate effect estimates, we run this model twice, changing only the feature used to represent masks; the priors and functional form are kept the same. Our model shares information across all countries to produce a statistically robust estimate, and thus measures the international mass wearing and mandate effects.

Figure 3A shows the effects we infer for wearing and mandates in the form of percentage reductions in *R*. We find that the difference between zero mask wearing and 100% self-reported mask-wearing (most or all of the time) corresponds to a 24.6% [6%, 43%] reduction in transmission. For mandates we see no reduction: 0.0% [−8.8%, 8.2%]. A more comparable measure is the probability of a positive reduction: for wearing this is 99%, while for mandates it is 46%. Together, these results suggest that mask-wearing is associated with a notable reduction in SARS-CoV-2 transmission, while analysis with mask mandate data yields no reduction.

**Fig. 3.**
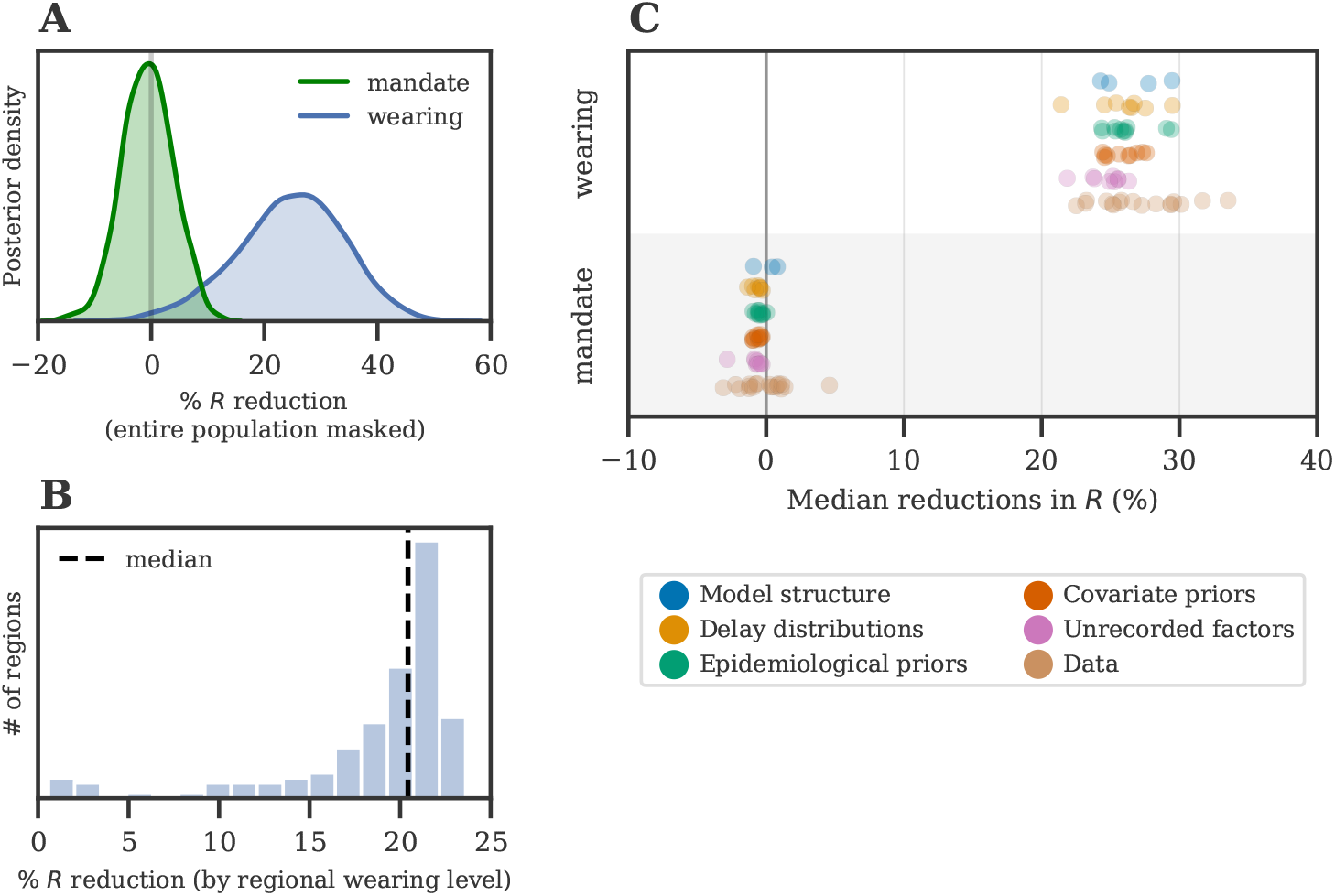
(A) Posterior reduction in *R* using wearing data (blue) and mandate data (green). Wearing posterior represents the % reduction in *R* when wearing increases from 0% to 100%. (B) Actualised reductions in transmission due to mask-wearing. Each reduction is the posterior median effect given the observed median level of wearing for each region, in this window. Dashed line = international median. (C) Estimates over 123 sensitivity experiments; each dot is the median under a different experimental condition. (‘Wearing’ denotes the 0-100% effect.)

Figure 3B shows the distribution of mask-wearing effects across the regions we study, using the *observed* median wearing percentage in each region. In this window of analysis, we infer a median reduction in transmission of 20.8% [2.7%, 23.2%].

All code and data used are available via Github: https://github.com/g-leech/masks_v_mandates.

## ROBUSTNESS

Results that are sensitive to alternative plausible modelling assumptions offer only weak evidence and pose a risk of misinforming policy decisions. As such, we verify the robustness of our results by performing 123 experiments across 22 sensitivity analyses (Table 1). Figure 3C shows how the median effect of wearing or mandates changes as we vary epidemiological priors, delay distributions, covariate effect priors, the model structure, and the data. Each point in Figure 3C is the median effect of a different experimental condition. Our results are robust to these changes−95% of the median reductions fall between 22.7% and 31.3%.

**Table 1.**
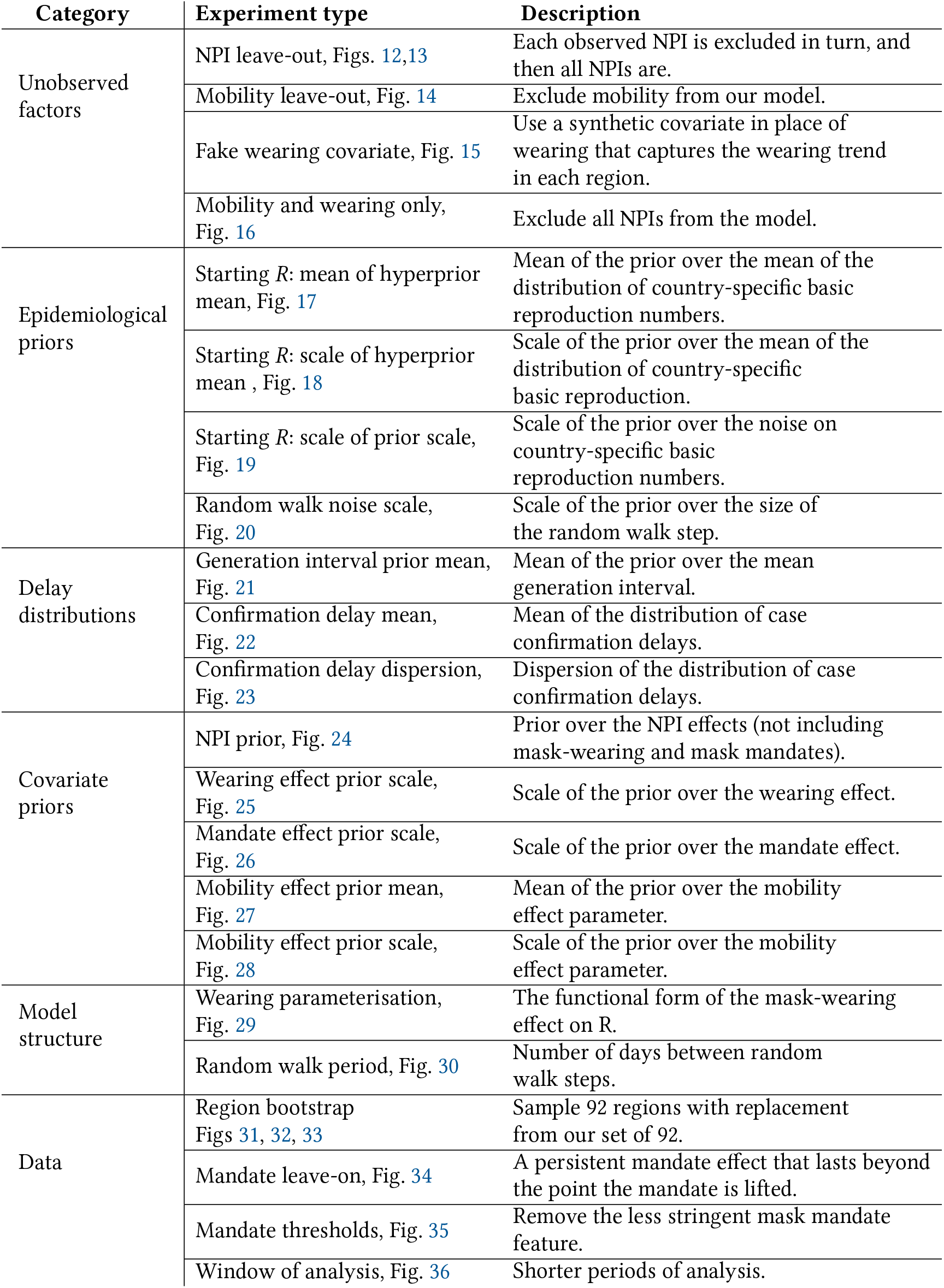
Experiments in our sensitivity analysis

| Category | Experiment type | Description |
| --- | --- | --- |
| Unobserved factors | NPI leave-out, Figs. 12,13 | Each observed NPI is excluded in turn, and then all NPIs are. |
|  | Mobility leave-out, Fig. 14 | Exclude mobility from our model. |
|  | Fake wearing covariate, Fig. 15 | Use a synthetic covariate in place of wearing that captures the wearing trend in each region. |
|  | Mobility and wearing only, Fig. 16 | Exclude all NPIs from the model. |
| Epidemiological priors | Starting $R$: mean of hyperprior mean, Fig. 17 | Mean of the prior over the mean of the distribution of country-specific basic reproduction numbers. |
| | Starting $R$: scale of hyperprior mean, Fig. 18 | Scale of the prior over the mean of the distribution of country-specific basic reproduction. |
| | Starting $R$: scale of prior scale, Fig. 19 | Scale of the prior over the noise on country-specific basic reproduction numbers. |
|  | Random walk noise scale, Fig. 20 | Scale of the prior over the size of the random walk step. |
| Delay distributions | Generation interval prior mean, Fig. 21 | Mean of the prior over the mean generation interval. |
|  | Confirmation delay mean, Fig. 22 | Mean of the distribution of case confirmation delays. |
|  | Confirmation delay dispersion, Fig. 23 | Dispersion of the distribution of case confirmation delays. |
| Covariate priors | NPI prior, Fig. 24 | Prior over the NPI effects (not including mask-wearing and mask mandates). |
|  | Wearing effect prior scale, Fig. 25 | Scale of the prior over the wearing effect. |
|  | Mandate effect prior scale, Fig. 26 | Scale of the prior over the mandate effect. |
|  | Mobility effect prior mean, Fig. 27 | Mean of the prior over the mobility effect parameter. |
|  | Mobility effect prior scale, Fig. 28 | Scale of the prior over the mobility effect parameter. |
| Model structure | Wearing parameterisation, Fig. 29 | The functional form of the mask-wearing effect on $R$. |
|  | Random walk period, Fig. 30 | Number of days between random walk steps. |
| Data | Region bootstrap Figs 31, 32, 33 | Sample 92 regions with replacement from our set of 92. |
|  | Mandate leave-on, Fig. 34 | A persistent mandate effect that lasts beyond the point the mandate is lifted. |
|  | Mandate thresholds, Fig. 35 | Remove the less stringent mask mandate feature. |
|  | Window of analysis, Fig. 36 | Shorter periods of analysis. |

However, as this study is observational rather than experimental, caution is necessary when making causal interpretations. Unobserved factors may influence *R*, and if their timing coincides with the timing of mask-wearing and mandates, reductions in *R* from unobserved factors may be wrongly attributed to mask-wearing or mandates [35]−our observed factors will be confounded. For instance, other protective behaviours may potentially confound our estimates [1, 15]. We investigate the susceptibility of our results to such confounding in four sensitivity analyses. In the first three (Figures 12, 14, 16), we assess how much estimates change when we exclude previously observed factors: we exclude each NPI in turn, all NPIs at the same time, and the mobility covariate. The small difference between our adjusted and unadjusted estimates suggests that, unless the confounding from unobserved factors greatly exceeds the confounding from our previously-observed factors (that is, NPIs and mobility), our results are unlikely to be meaningfully affected by confounding [36]. Lastly: over our window of analysis, mask-wearing increases while transmission decreases (in many regions). Our final analysis aims to assess whether this correlation is a spurious contributor to the substantial apparent wearing effect. We test this hypothesis by creating a fake wearing variable for each region. Each variable has the same start and end wearing value as the true wearing percentage and linearly interpolates between these values to capture the trend in wearing in that region. We infer a small and uncertain effect for the fake wearing variable 7.6% [–20.2%, 30.0%] (see Figure 15). This implies that the wearing effect we infer does not rely solely on the correlation between transmission and the overall wearing trend in this period.

## DISCUSSION

We find that mask-wearing is associated with a notable reduction in SARS-CoV-2 transmission. Moreover, using data on mandates fails to infer any reduction in transmission. Our results suggest that national (and US state-level) mandate data are insufficient to model the effect of mass mask-wearing. Figure 2 illustrates several ways mandates can fail to correlate with wearing: South Korea’s mandate came after voluntary wearing had already plateaued at 94%; conversely, in the Netherlands and Switzerland, few people were wearing masks, even three weeks into the mandate period; finally, in the Czech Republic, wearing eventually increased, but only long after the mandate was implemented.

### Against mandate *data*, not mandates

In our window, national mandates correspond to an average 8.3% increase in the number of people who say that they are likely to wear masks most or all of the time in public spaces; however, this may underestimate the effect of mandates on wearing. This could be the case if mandates encourage people to wear masks in public *all* the time instead of *most* of the time, or if there is large sub-national heterogeneity in mandate timing and wearing uptake.

Inferring mandate effects is also difficult with currently available data. We model the effect of mandates as an instantaneous change in the reproduction number. This does not capture changes in wearing behaviour following the announcement of a mandate but before its enforcement [21]. Nor does it account for gradual change in behaviour after the implementation of a mandate.

### Heterogeneity

The variation in results discussed in the Introduction is in part due to not controlling for mask properties and wearing behaviour. These include mask quality [37]; mask fit [37]; the *venue* of wearing (e.g. in shops, schools, or public transport) [37]; mask reuse [38]; risk compensation [39]; and cultural norms [16, 37, 39]. More research into these factors is required to further reduce our uncertainty about mask-wearing effects. We estimate the effect of mass mask-wearing, averaging over mask properties and behaviour. Given that, in this window, most masks in use were the least effective types (cloth or otherwise unrated masks) [1, 13, 14, 38, 40], the effectiveness of mass wearing is likely stronger than we estimate. Finally, we report the average international effect of mandates and do not rule out their effectiveness in particular contexts; for example, strong correlations between mandates and wearing were observed in Ireland (Figure 2) and in Germany (the April 2020 local mask mandates [15, 21]). Our results should be adjusted to local circumstances by public health experts.

### Window of analysis

Our results are based on the period from May to September 2020. While we find similar results for different (shorter) windows of analysis (Figure 36), mass wearing effectiveness will likely differ with larger changes in circumstances. In particular, our period has features that may not characterise other settings: most regions began with NPIs already active (besides mandates); public behaviour had already changed following the formal and informal instructions of the first wave; summer months are thought to have lower transmission [41, 42]; and a tiered regional approach to containment was not yet implemented in most regions. However, a short window implicitly holds many factors constant. This is useful for internal validity: when estimating a specific quantity such as the effects of mask-wearing, a short window reduces the scope for distribution shift and unobserved confounders.

### Operationalising mask-wearing

Mask-wearing surveys are still a proxy for actual wearing behaviour, and social desirability bias in survey responses may inflate wearing estimates [43]. In a Kenyan study, the disparity between self-reported wearing and observed wearing was 77% [44]−though this survey was not anonymous, which may have lead to more over-reporting than anonymous surveys such as COVIDNearYou−SurveyMonkey. If data sources over-estimate mask-wearing, then our estimate for the effect of 100% of people wearing masks (most or all of the time) will actually correspond to the effect of less than 100% of people wearing masks. Consequently, we would expect the true effect of 100% of people mask-wearing to be larger than we estimate, in proportion to the amount of over-reporting. Further, the operational definition of ‘mask-wearing’ used in the UMD survey is not stringent: it can be applied both to a person who wears a cloth mask, only on public transport, slightly more than half of the time; and to a person who always wears an N95 respirator when outside their home [7]. This implies that there is scope for more and better mask-wearing, even in regions reporting extremely high levels of wearing in our data.

### Endogeneity of interventions

One concern for observational NPI studies is endogeneity: when cases are rising, people are more likely to voluntarily mask and governments are more likely to mandate wearing [45]. However, in our window, the correlation between new cases and mask-wearing percentage is low, *ρ* = 0.05, which limits the scope of this concern.

### Conclusion

At a time where mask-wearing is decreasing and mask mandates are being lifted, we find that mask-wearing is associated with a notable reduction in transmission, and that factors other than mandates must have contributed to the worldwide uptake of mask-wearing in 2020. This presents a difficulty for policy-makers: if wearing works but mandates are not strongly associated with wearing, what other levers are available? Some options include free mask distribution, domestic supply guarantees, fit training, mask quality guidelines, targeted mandates by venue, and openness about the benefits of masks [2, 46].

## METHODS

All data and code used can be downloaded via: https://github.com/g-leech/masks_v_mandates. The preprocessing is derived from [29].

### Data

Our analysis is on the national (or US state) level, since this is the finest resolution available for all countries in the OxCGRT NPI dataset. Table 2 summarises the modelling set, and Figure 4 shows its component datasets.

**Table 2.** Modelling data summary

|  |  |
| --- | --- |
| **Regions** | 92 (55 countries + 37 US states) |
| **Period** | 1st May 2020 - 1st Sep 2020 |
| **Modelling data points** | 13,248 days across all regions |
| **Mask-wearing data points** | 19.97m (UMD) + 558,670 (COVIDNearYou) |
| **Data validation** | Manual correction of reporting errors;<br>filtering out non-epidemic regions;<br>validation against external sources |

**Fig. 4.**
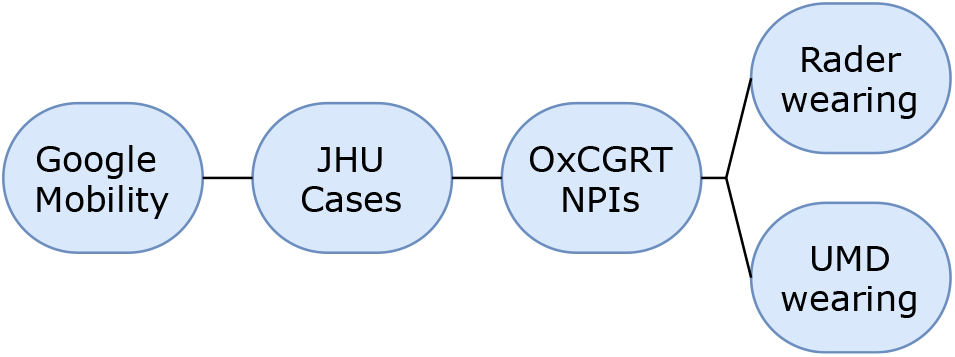
The components of our modelling set. ‘Google’ [48]; ‘JHU’ [47]; ‘OxCGRT’ [32]; ‘UMD’ [7]; ‘Rader’ [25].

The beginning of our window of analysis is determined by our datasets: the UMD project begins reporting in late April 2020 [7]. We end on the 1st September 2020, at the beginning of the second wave, a period in which national NPIs fragment into regional responses, making national analyses less informative [19].

Daily national estimates of mask-wearing are derived from the University of Maryland (UMD) / Facebook COVID-19 World Symptoms Survey [7], which randomly samples from all active Facebook users, and which post-stratifies to correct for nonresponse bias and demographic imbalance [33]. The mean number of individual responses per region-day is 1131. UMD does not cover the US, so we supplement this dataset with the US data of [25], which in our window represents n=558,670 responses.

Daily confirmed COVID-19 cases are drawn from the Johns Hopkins CSSE COVID-19 Data Repository, which collates official statistics from around the world [47].

We use the Google COVID-19 Community Mobility Reports to index mobility changes in each region [48].

See Appendix A for full data details, including preprocessing steps and country selection.

### Model

We develop a hierarchical Bayesian model based on prior work [19, 29, 49] to infer the effectiveness of mask wearing and mask mandates on COVID-19 transmission. We use the number of reported cases in each country to infer the number of later-ascertained infections on each day. Given the dynamics of daily, later-ascertained infections in each region over time, we infer the instantaneous reproduction number *R*_*t*_. Finally, the covariate effects are estimated by relating the *R*_*t*_ to the observed level of each covariate. Figure 5 shows the model in schematic form. The Bayesian approach allows us to explicitly model sources of uncertainty, such as the values of epidemiological parameters, which are known with uncertainty. We proceed by outlining the inputs of our model.

**Fig. 5.**
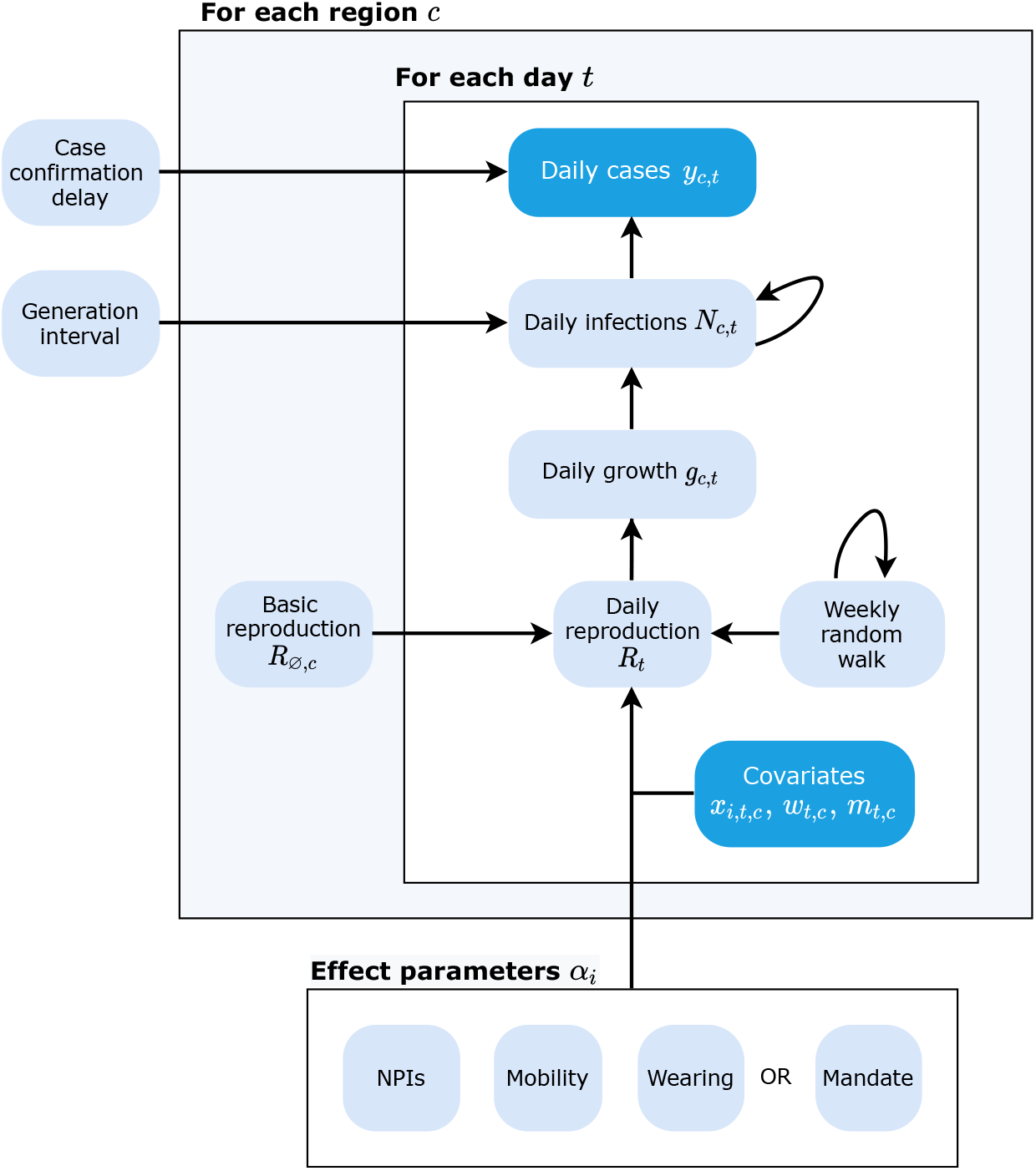
Schematic of our model. Observed nodes in dark blue, latent nodes light blue. The target of our analysis (bottom) is *α*_*i*_. On each day *t*, region *c*’s reproduction number *R*_*t*_ depends on: 1) the starting reproduction number *R*_∅,*c*_, 2) the NPIs active in *c*, 3) the mobility level, 4) either the wearing level or the mandate indicator, and 5) a location-specific weekly random walk. The resulting *R*_*t*_ estimate (in the form of a growth rate) is used to compute the latent daily infections *N*_*t*_, given the distributions over the generation interval and the previous infection count. Finally, the expected number of daily confirmed cases (*y*_*t*_) are computed using *N*_*t*_ and the distribution over the delay until case confirmation.

### Notation

We use *c* to denote the country/region in question, and *t* to index time. *t* = 0 corresponds to May 1st, 2020. NPIs are indexed by *i*.

### Inputs

- **Non-pharmaceutical interventions (NPIs)**: *x*_*i,t,c*_ ∈ {0, 1}. *x*_*i,t,c*_ = 1 if NPI *i* is active at time *t* in region *c*; otherwise, *x*_*i,t,c*_ = 0.
- **NPI reopenings**: Across our regions, there are NPIs that were active at the start of our period. We treat these NPIs, in the relevant regions, as ‘reopening’ NPIs. If NPI *i* is active in region *c* at *t* = 0 (i.e. we have *x*_*i*,0,*c*_ = 1), we subtract 1 from the feature to form 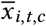. Therefore, at the start of the window, 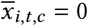 and the effect of the NPI is absorbed into *R*_*ϕ,c*_. When the NPI lifts, we would have 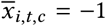, reflecting that NPI lifting has the opposite effect to NPI closing, which is denoted as *x*_*i,t,c*_ = 1. As such, we can more easily set a prior over *R*_∅,*c*_ (see below).

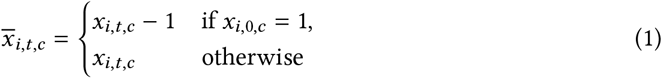
- **Mask mandate**: We have two mandate covariates: *x*_ma1,*t,c*_ and *x*_ma2,*t,c*_. The first covariate, *x*_ma1,*t,c*_, represents whether masks were ‘*required in some or all shared spaces, outside the home with other people present, or some situations when social distancing not possible*’ (field H6 from OxCGRT, level 2 [32]). The second covariate, *x*_ma2,*t,c*_, has the same conditions, but masking is required in all shared spaces (field H6 from OxCGRT, level 3 or higher). For each mandate type, *x*_ma,*t,c*_ = 1 if a mask mandate corresponding to the description above is active at time *t* in region *c*; otherwise, *x*_ma,*t,c*_ = 0. *x*_ma1,*t,c*_ = 1 whenever *x*_ma2,*t,c*_ = 1, so the correct interpretation of the effect associated with *x*_ma2,*t,c*_ is the *additional* effect of mandating masks in all shared spaces, given that mask mandates were already required in some shared spaces.
- **Mask wearing**: The percentage of people in each region that self-report as likely to/always wear masks in public, *w*_*t,c*_ ∈ [0, 1].
- **Mobility**: Reduction in mobility relative to 2019 levels, *m*_*t,c*_ ∈ [−∞, 1], represented as a multiplicative factor. *m*_*t,c*_ = 1 represents a 100% decrease in mobility while *m*_*t,c*_ = 0 represents no change from 2019 level.
- **Cases**: New confirmed cases observed on day *t*: *y*_*t,c*_.

In the following sections, we introduce several variables without explicitly defining them. They are defined in the section on Prior Distributions below.

### Infection Model

The instantaneous reproduction number *R*_*t,c*_ is the expected number of infections that would arise from each infection at time *t* in region *c*, all else equal. We model *R*_*t*_ as a product of the several terms: (i) the regional starting reproduction number *R*_∅,*c*_; (ii) a product of our effect estimates for that region-day for each of the reopening NPIs *X*_*t,c*_, mask-wearing *W*_*t,c*_ *or* mask mandates Ma_*t,c*_ (mask-wearing is shown), and mobility *M* (*m*_*c,t*_)^−^; (iii) a weekly latent random walk per region *z*_*t,c*_.

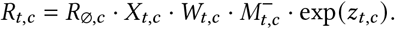

We will now discuss each of these terms in turn:

#### Latent reproduction number

The latent, unobserved reproduction number in region *c* at *t* = 0, assuming no mask-wearing and no active mask mandates, is represented by *R*_∅,*c*_.

#### NPIs

We assume that the introduction or lifting of an NPI leads to an instantaneous, multiplicative change in transmission. Each NPI contributes 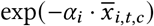 to *R*_*t,c*_. Note that this also works for reopening NPIs−if the NPI effect (*α*_*i*_) is positive, a reopening 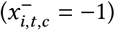 increases *R*:

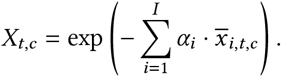

#### Mask mandates

In the mandate model, *W*_*t,c*_ is replaced with Ma_*t,c*_ = exp (−*α*_ma1_ *x*_ma1,*t,c*_) · exp (−*α*_ma2_ *x*_ma2,*t,c*_).

#### Mask-wearing

*W*_*t,c*_ = exp (−*α*_*w*_*w*_*t,c*_). We use the exponential form in our base model because it is consistent with the form of the mandate effect on *R*. However, we test the sensitivity of our results to two alternative mask-wearing parameterisations and find similar results (see Appendix C).

#### Mobility

We parameterise the Google mobility data as in [50]:

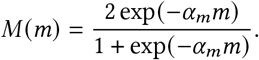

At 2019 levels of mobility (*m* = 0), the multiplicative factor *M* (*m*) = 1, leading to no effect on *R*_*t*_. To set a principled prior for *R*_∅,*c*_, we zero-center the mobility by subtracting the initial level (see the section on Prior Distributions):

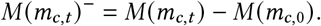

#### Random walk

The weekly random walk is computed as:

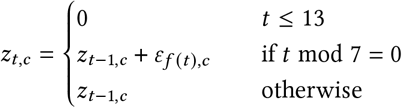

where *f* (*t*) = ⌊(*t* − 14)/7⌋ and *ε* ∼ `Normal` (0, *σ*_RW_). The random walk starts after 2 weeks to avoid unidentifiability between *R*_∅,*c*_ and the random walk terms at the beginning of the period.

Following [51], the resulting *R*_*t*_ estimate is then transformed to daily growth using the generation interval distribution, which describes the time between success infection events in a transmission chain. *N*_*t,c*_ represents daily infections that are later ascertained, and we have *N*_*t,c*_ = *g*_*t*−1,*c*_ · *N*_*t*−1,*c*_ i.e., we multiply the infections on the previous day by the daily growth rate. Then, given an initial (latent) infection count, we have:

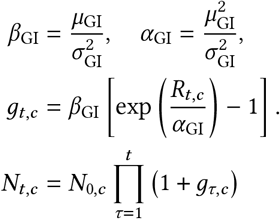

### Observation Model

Infections at time *t* are only observed as reported cases after a delay. Therefore, we convolve the later-ascertained cases with a delay vector to produce 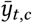, which is the expected number of reported cases on day *t* in region *c*.

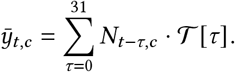

The forward-delay vector 𝒯 (defined in Prior Distributions, below) defines the delay between the two quantities. Finally, the observed number of reported cases, *y*_*t,c*_, follows a Negative Binomial distribution:

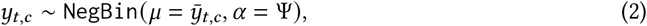

where Ψ is the case-reporting overdispersion parameter (see below).

### Prior Distributions

We place prior and hyperprior^2^ distributions over several parameters. Our Bayesian approach not only captures uncertainty in unknown parameters, but allows our beliefs about certain parameters to be adjusted if warranted by the data. We now detail the priors we use in this work.

- **Region-specific** *R*_∅_: *R*_∅,*c*_ ∼ `Normal`(*μ*_*R*_, *σ*_*R*_);.
- *R*_∅_ **hyperpriors**: The Epidemic Forecasting group [52] produces estimates for *R*_*t,c*_ using methodology from [53]. The empirical mean and variability of these estimates across our regions at the start of our period is *μ* = 1.07, *σ* = 0.32. We use these estimates to initialise our hyperpriors over the mean and variability of *R*_∅,*c*_: *μ*_*R*_ = `TruncatedNormal`(*μ* = 1.07, *σ* = 0.2, `lower`= 0.1), *σ*_*R*_ = `HalfNormal`(*σ* = 0.4). The median of *σ*_*R*_ under this prior is 0.32.
- **NPI effect:** *α*_*i*_ ∼ `AsymmetricLaplace`(*m* = 0, *κ* = 0.5, *λ* = 30), following [29]. *m* is the location, *κ* is the asymmetry, and *λ* is the scale. This prior places 80% of its mass on positive NPI effects (i.e. on reductions of *R*).
- **Wearing effect**: *α*_*w*_ ∼ `Normal`(*μ* = 0, *σ* = 0.4). Unlike the NPIs above, the prior for wearing has equal mass on positive and negative effects. This uninformative choice reflects past uncertainty about the efficacy of mask-wearing.
- **Mandate effect**: *α*_ma_ ∼ `Normal`(*μ* = 0, *σ* = 0.08). The wearing prior reflects our prior beliefs about the effect of going from 0-100% of people likely to wear masks. But in our window, the range of *w*_*t,c*_ averages only ∼20% across our regions. Accordingly, we choose a prior for the effect of mandates that has 1/5th of the prior predictive effect as the wearing prior. In our sensitivity analysis we modify the scale of this prior to match that of the wearing prior (among other values), and find very similar results.
- **Mobility effect**: *α*_*m*_ ∼ `Normal`(*μ* = 1.704, *σ* = 0.44). Mobility prior values are derived from the ‘overall average mobility’ estimate in [50]. Note that each *α* above is not a direct reduction in *R*; they are transformed into a reduction via a specific functional form (see above).
- **Initial infection counts:** Initialised with the empirical median new confirmed cases of the first day of our window, 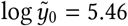.

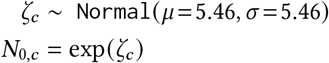
- **Random walk noise scale**, chosen as in [19]

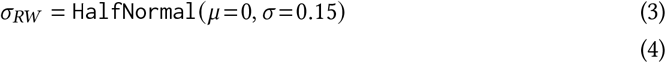
- **Generation interval distribution** [54, 55]:

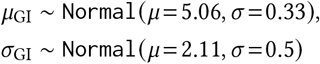
- **Time from infection to case confirmation** 𝒯 [29, 55–57]: The delay between infection and case confirmation is distributed as

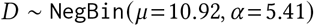 We produce a forward-delay vector

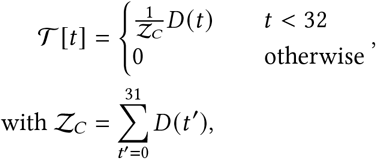

i.e., a negative binomial distribution, truncated at 31 days and normalised. Note that the Negative Binomial *α* parameter denotes the dispersion, not the variance, 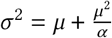.
- **Observation noise dispersion**, chosen as in [29]

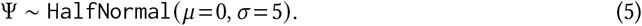

## Data Availability

All data can be downloaded with the instructions on the Github repo:
https://github.com/g-leech/masks_v_mandates

## ACKNOWLEDGMENTS

We thank Swapnil Mishra for cloud infrastructure and moral support; we thank Tomáš Gavenčiak for help debugging and plotting. We thank Jan Kulveit for strategising, and Joshua Monrad for preternatural style advice.

## SUPPLEMENTARY INFORMATION

### A DATA

#### JHU CSSE cases database

We take daily confirmed case counts from the Johns Hopkins University Center for Systems Science and Engineering COVID-19 Global Cases dataset, which collates official statistics from hundreds of world regions.

Many countries fail to report case numbers over the weekend (or report weekly), which leads to spurious periodicity. In addition, severe reporting errors (day-to-day spikes of 1000% or troughs of less than 10% in countries with hundreds or thousands of daily cases) occur in 23 regions. We manually mask these errors (Table 3), preventing the model from learning from those days.

**Table 3.** Dates of reporting errors in the JHU case data

|  |  |
| --- | --- |
| **Costa Rica** | 2020-09-20, 2020-09-21 |
| **Ethiopia** | 2020-06-30, 2020-07-01 to 2020-07-08 |
| **Guatemala** | 2020-07-18 |
| **Lebanon** | 2020-08-04, 2020-08-05 |
| **Libya** | 2020-08-23, 2020-08-24 |
| **Michigan** | 2020-08-21, 2020-08-22, 2020-08-29, 2020-08-30 |
| **United Kingdom** | 2020-07-01, 2020-07-02 |
| **Honduras** | 2020-05-20 to 2020-05-24, 2020-05-29, 2020-05-30 |
| **Netherlands** | 2020-08-11, 2020-08-12 |
| **Panama** | 2020-06-14, 2020-06-15 |
| **Singapore** | 2020-08-05 |
| **Serbia** | 2020-07-25, 2020-07-26 |
| **Alabama** | 2020-06-27, 2020-06-28 |
| **Arizona** | 2020-06-29 |
| **Colorado** | 2020-09-04, 2020-09-05 |
| **Delaware** | 2020-05-23, 2020-05-24 |
| **Minnesota** | 2020-07-04 |
| **New Mexico** | 2020-05-23, 2020-05-24 |
| **Oregon** | 2020-06-06 to 2020-06-08, 2020-06-13, 2020-06-14 |
| **South Carolina** | 2020-06-03 to 2020-06-07 |
| **Washington** | 2020-05-23, 2020-05-24 |
| **Wisconsin** | 2020-08-18, 2020-08-19 |
| **Iowa** | 2020-08-27 |

#### The OxCGRT NPI database

We take NPI data from the Oxford COVID-19 Government Response Tracker, which collects data at the national-level and US state-level [32]. From these we select the ‘containment’ policies, i.e. direct attempts to reduce transmission.

Importantly, OxCGRT cannot be used for national modelling without imputation. OxCGRT reports only one value per country-day, even if policies differ between regions. The dataset reports the maximum stringency of each NPI, whether or not this is implemented in all regions. This leads to the national stringency value being “hidden” behind the highest regional value, where any region has stronger measures. As a result, when a policy is strengthened in only part of a country, we impute the previous national value.

We process the NPI data as follows:

- We filter to rows with national coverage (that is, ‘Flag’ columns = 1).
- We threshold the ordinal values as in (4), creating a feature for the first mandatory level of each policy and additional features for higher levels of school closing, workplace closing and restrictions on gatherings. This yields 10 NPI features.
- When a policy is strengthened in only part of a country, we impute the previous national value.

#### UMD / Facebook wearing dataset

We use the University of Maryland Centre for Geospatial Information Science—Facebook Research survey as our main source of daily, self-reported wearing data [7, 33]. This is by far the largest-scale survey of COVID mask-wearing (with 19.97 million individual responses in our window, or 1,500 individual responses per region-day). The survey uses stratified random sampling of all active Facebook users to ensure demographic balance in each region, and also guarantees at most one response per month per Facebook user.

An alternative survey, the Imperial College London—YouGov COVID-19 Behaviour Tracker [58] is one hundred times smaller than UMD, uses nonrandom sampling, and has most days missing, and is as such less suitable for modelling.

#### The COVIDNearYou / SurveyMonkey United States wearing dataset

The UMD dataset does not include US wearing data, while the respective CMU / Facebook US survey [59] does not begin reporting until after our window of analysis. We supplement UMD with data from Rader *et al*. [25].

The Rader data are individual survey responses on a reverse Likert scale, weighted to correct for demographic imbalance in the sample. To convert this to the UMD scale, we take the mean of the grocery shopping and workplace features, threshold at ≤ 2 (likely or very likely) and take the percentage of rows in each state passing this threshold, and smooth over a 7 day window. This results in a percentage-wearing feature which is within 1% of the Facebook US data [59] for the period where the two datasets overlap.

#### Google Mobility Index

We use the Google COVID-19 Community Mobility Reports to index mobility changes in each region [48]. We form a single feature by averaging the indoor public components (retail and recreation, grocery and pharmacy, transit, and workplaces). We parameterise mobility similarly to Unwin *et al*. [50].

#### Instantaneous reproduction number estimates

To validate our model estimates, and for the initialisation of *R*_0_, we use country *R*_*t*_ estimates from the Epidemic Forecasting group [52]. The estimates are calculated using a nonparametric approach from [53]. US state-level estimates are taken from https://rt.live/.

#### Country selection

The OxCGRT dataset has 184 countries, or 235 counting US territories. 81 countries are missing from the UMD wearing data, and are thus dropped when joining to OxCGRT. We manually drop 32 countries with frequent extreme periodicity in case reporting, 16 countries that have fewer than 5000 cumulative cases in our window, 10 countries not contained in the Google Mobility dataset, and 4 countries that are missing more than 3 consecutive weeks of wearing data. Included countries are shown in Table 5.

**Table 4.** OxCGRT NPI features and our threshold choices

| **Original feature name** | **Original scale** | **Cutoff** |
| --- | --- | --- |
| C1_School closing | Ordinal (0-3) | 2,3 |
| C2_Workplace closing | Ordinal (0-3) | 2,3 |
| C3_Cancel public events | Ordinal (0-2) | Obsoleted |
| C4_Restrictions on gatherings | Ordinal (0-4) | 2, 3, 4 |
| C6_Stay at home requirements | Ordinal (0-3) | 2 |
| C7_Restrictions on internal movement | Ordinal (0-2) | 2 |
| H6_Facial Coverings | Ordinal (0-4) | 2, 3 |
| *_Flag | Binary | - |

**Table 5.** Regions included in the analysis, by continent

| Continent | Region |
| --- | --- |
| Asia | Bangladesh, India, Indonesia, Iraq, Israel, Japan, Lebanon, Nepal, Philippines, Saudi Arabia, South Korea, Singapore, United Arab Emirates |
| Europe | Austria, Belarus, Croatia, Czech Republic, Germany, Greece, Hungary, Ireland, Italy, Moldova, Netherlands, Norway, Poland, Portugal, Romania, Russia, Sweden, Turkey, Ukraine, Switzerland, United Kingdom |
| Africa | Egypt, Morocco, Libya, Kenya, Nigeria, South Africa |
| South & Central America | Argentina, Bolivia, Brazil, Colombia, Costa Rica, Dominican Republic, El Salvador, Guatemala, Honduras Mexico, Panama, Paraguay, Venezuela |
| North America | Canada, Alaska, Alabama, Arkansas, Arizona, California, Colorado, Delaware, Florida, Georgia, Hawaii, Iowa, Illinois, Indiana, Massachusetts, Maryland, Michigan, Minnesota, Missouri, Montana, North Carolina, North Dakota, Nebraska, New Jersey, New Mexico, Nevada, New York, Ohio, Oklahoma, Oregon, Pennsylvania, South Carolina, South Dakota, Texas, Utah, Virginia, Washington, Wisconsin |
| Oceania | Australia |

#### Mask-wearing and mask mandates in the first wave

The YouGov survey [58] begins in Jan 2020 for some locations, which enables us to check the mandate-wearing relationship in the first wave, at the time of the earliest mandates. Figure 6 displays the estimates against mandate date (including some countries with multiple mandates). The average reported level of mask-wearing in Jan 2020 was 32.7%. This increased to an average of 64.2% before the first national mandate implementations in March and April. There was an average post-mandate increase in wearing of 11%, similar to in our modelling set (an 8.3% post-mandate increase).

**Fig. 6.**
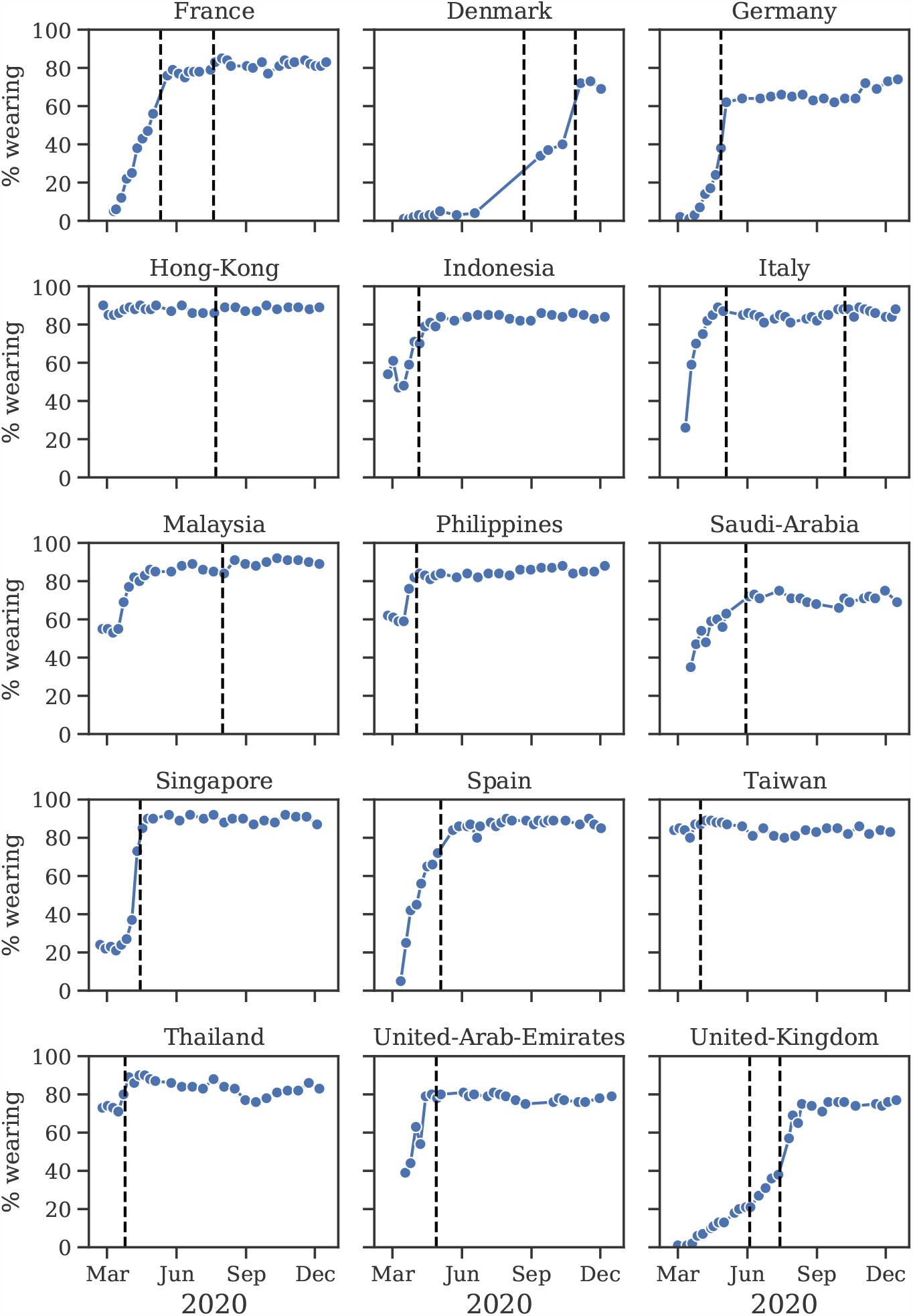
YouGov wearing estimates over time, with mandates as dashed lines [58]. These countries are those with both YouGov estimates and national mask mandates.

#### Mask recommendations

We follow past work in timing mandates with the beginning of the nominal legal enforcement of wearing. Our source of NPI data [32] also contains an indicator for whether a non-mandatory government recommendation to wear masks was in place. To see if this less stringent, but generally earlier, policy has stronger correlations with subsequent mask-wearing, we repeat the exploratory analysis from above. The correlation between wearing percentage and any form of recommendation or mandate is weaker than before, Spearman’s *ρ* = 0.235, *p* < 0.001, compared to the mandate correlation of 0.32.

### B MODEL OUTPUTS

#### MCMC statistics

We use PyMC3’s implementation of Hamiltonian Monte Carlo with the No-U-Turn sampler (NUTS) [60]. The following outputs result from running the default model with the wearing feature.

The Gelman-Rubin diagnostic 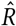 tests for convergence of the sampler. When 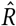 is close to 1 (i.e. < 1.01 [61]), the MCMC sampling algorithm is commonly considered to have converged [62]. Figure 7 (left) therefore suggests that our MCMC sampler has converged, and that our posterior may be used to draw valid inferences.

**Fig. 7.**
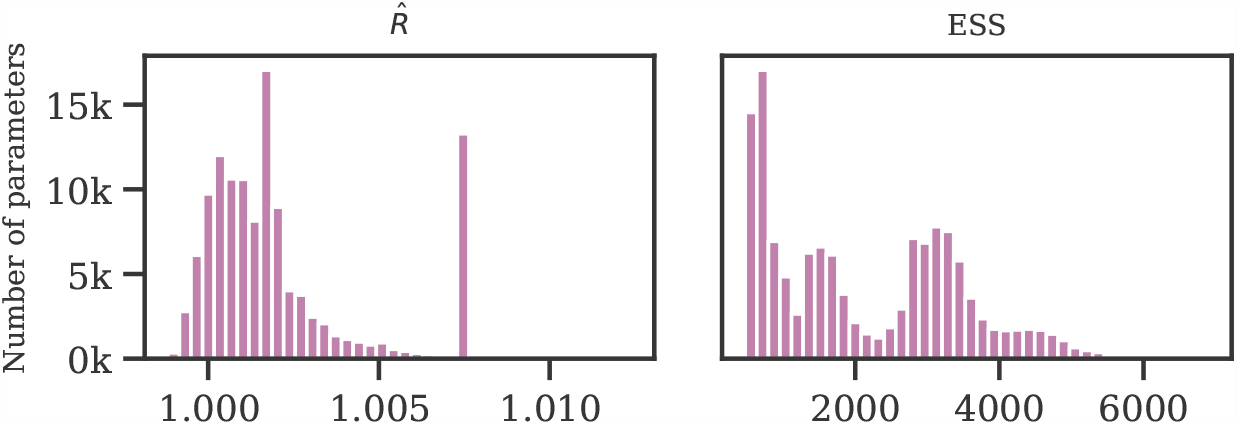
Gelman-Rubin 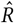 score (left) and effective sample size (right).

We used 1000 tuning samples and 500 posterior samples for each of 4 chains, giving 6000 samples in total. As shown in Figure 7 (right), the relative effective sample size exceeds 30% for the majority of parameters, indicating low autocorrelation.

#### Prior-posterior plots

Figure 8 displays the priors and posteriors for parameters of our model. The posteriors are sharp despite broad priors, which suggests that our data is informative about the parameters.

**Fig. 8.**
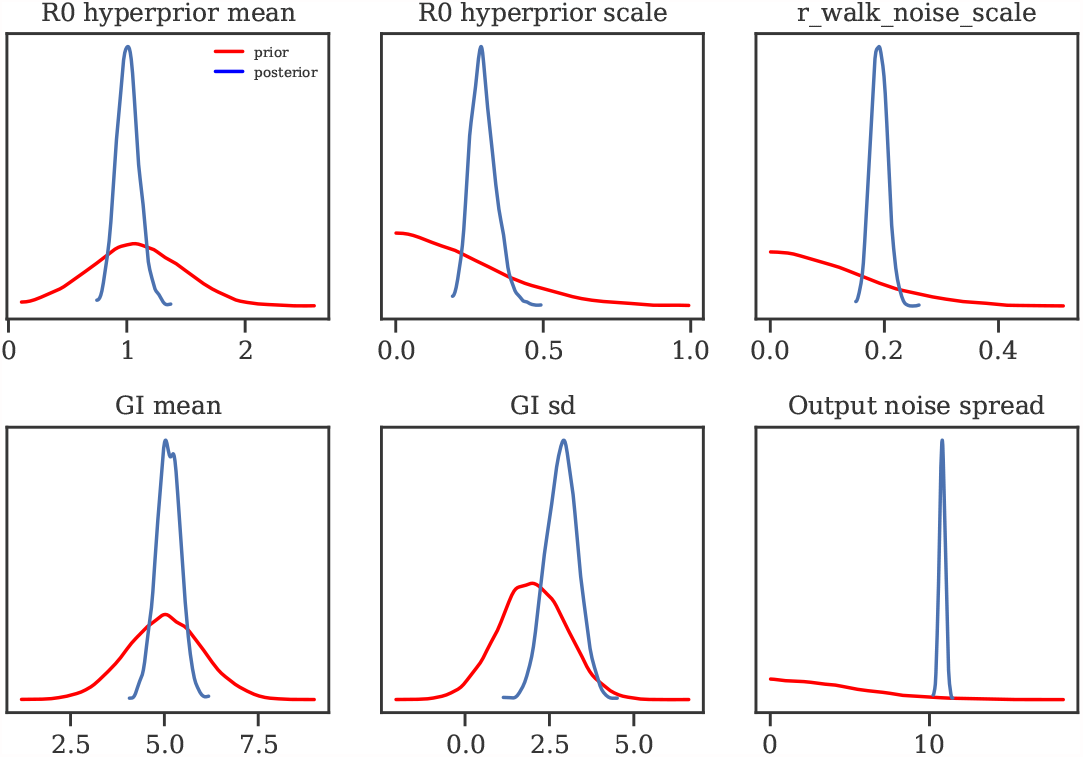
Priors vs posteriors for learned model parameters.

#### Posterior predictive distributions

Figure 9 displays predicted cases during and 3 weeks beyond our window of analysis. All 92 country panels can be found on [Github](https://github.com/g-leech/masks_v_mandates/blob/main/outputs/region_panels_wearing.pdf).

**Fig. 9.**
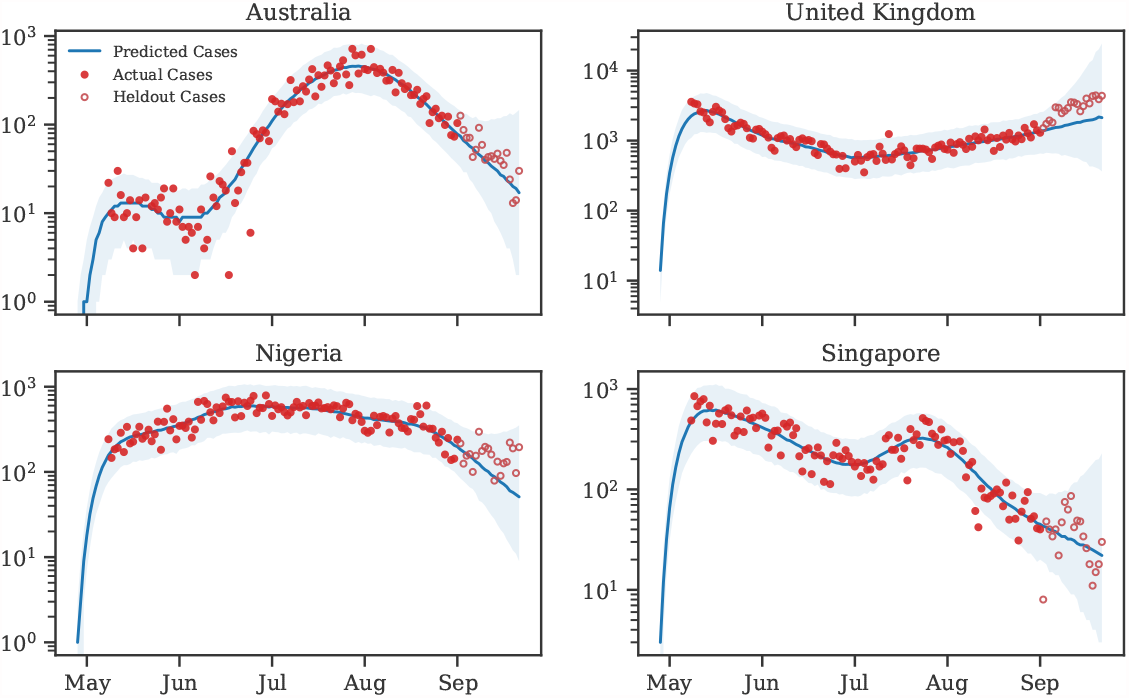
Predictive curves from selected regions. The last 20 data points are holdouts, unseen by the model.

#### Posterior correlations

Figure 10 shows the posterior correlations between the attributed R reductions for each modelled effect.

**Fig. 10.**
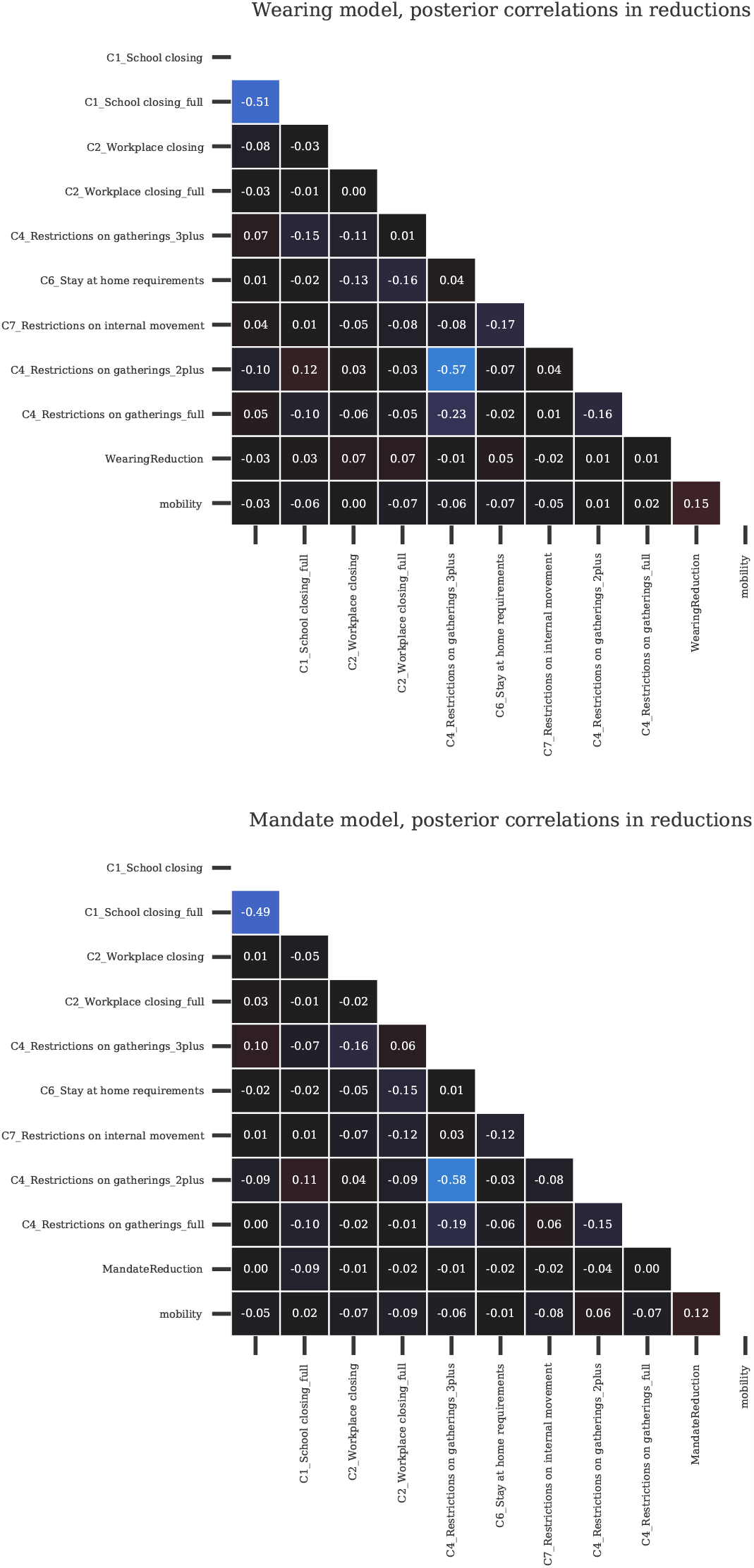
Posterior correlations between the covariate effects (reductions in R)

We can use these correlations to diagnose excessively strong collinearity in our data; collinearity would manifest as strong posterior correlations [63]. However, almost all of the pairwise correlations are −0.2 < *r* < 0.1, which indicates that collinearity is manageable in our dataset. Notable negative effect correlations exist between different levels of the same NPIs:

- Restrictions on gatherings < 100 people and Restrictions on gatherings < 1000 (−0.57);
- Restrictions on gatherings <10 and Restrictions on gatherings <100 (−0.23);
- School reopening (some schools) and School reopening (all schools) (−0.51);

All other pairwise covariate correlations have an absolute value less than 0.2.

#### Region panels

Figure 11 displays inferred *R*_*t*_ against covariate values for selected countries. All 92 country panels can be found on [Github](https://github.com/g-leech/masks_v_mandates/blob/main/outputs/region_panels_wearing.pdf).

**Fig. 11.**
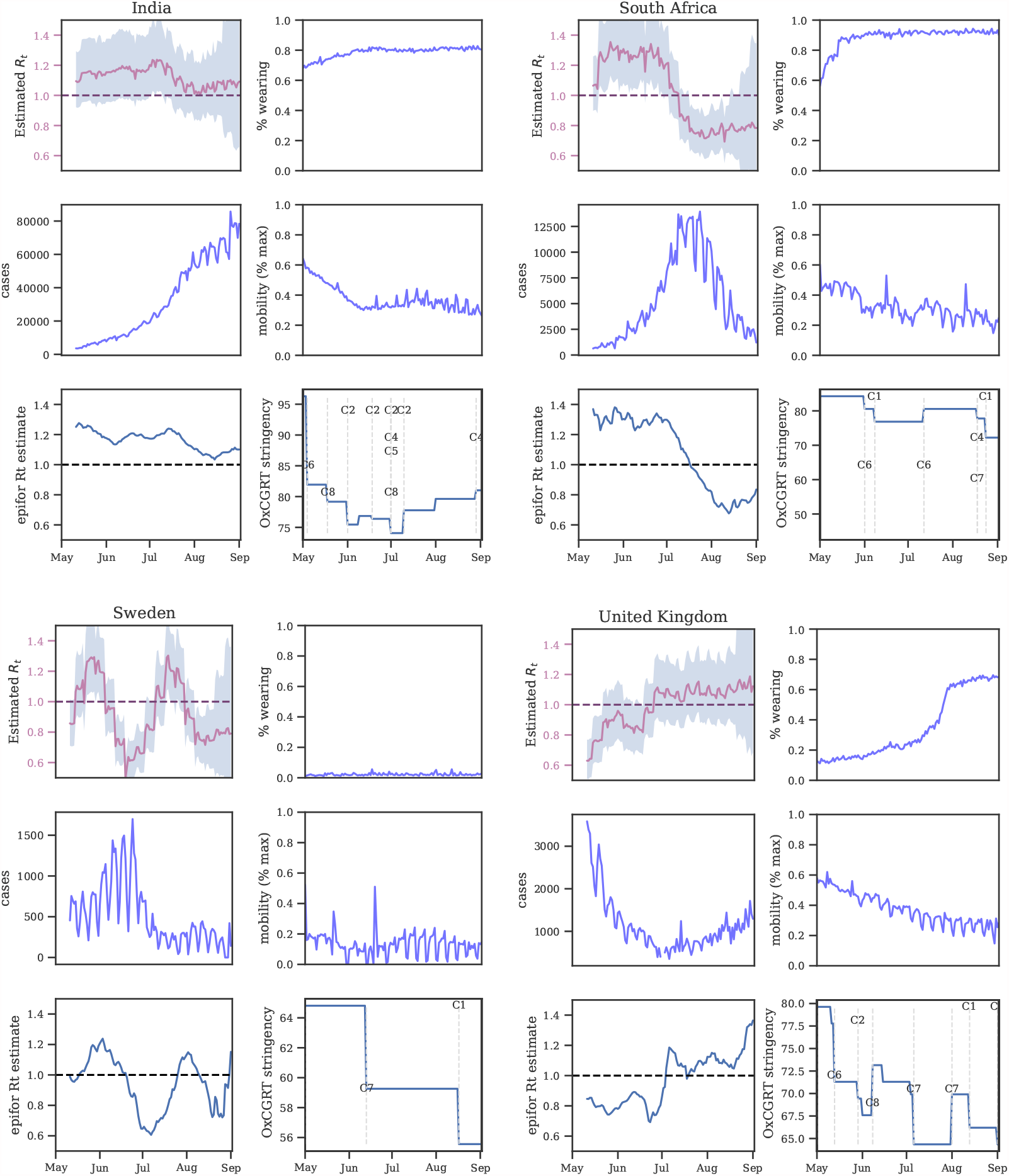
Summary plots of selected region covariates and *R*_*t*_ estimates, summer 2020. Top-left: instantaneous *R*_*t*_ from our model. Bottom-left: instantaneous *R*_*t*_ estimates from EpidemicForecasting [52]. Bottom-right: overall NPI stringency from OxCGRT [32]

### C SENSITIVITY ANALYSIS

Sensitivity analysis reveals the extent to which results depend on uncertain parameters and modelling choices, and can diagnose model misspecification and excessive collinearity [63]. We vary many of the components of our model and recompute the NPI effectiveness estimates. Overall, we perform 22 sensitivity analyses with 123 experimental conditions. Table 1 summarises our sensitivity analyses and their categories.

The effect sizes inferred for the other NPIs are smaller than in other work [19, 29, 49] because they measure a different effect: in this window, most regions begin with interventions active, and changes in NPI status are most often reopenings/lifting of bans. Such reopenings often result in an increase in transmission that is smaller in magnitude than the decrease in transmission from the initial policy implementation—for example, due to improved safety procedures [19].

#### C.1 Unobserved factors

Our data do not capture all of the government NPIs that were implemented, and we only measure two forms of voluntary behaviour change: mask wearing and mobility. Unobserved factors may influence *R*, and if their timing correlates with the timing of mask wearing or mandates, reductions in *R* from unobserved factors may be wrongly attributed to mask-wearing or mandates [35]—our observed factors will be confounded. For instance, observational estimates like ours are potentially confounded by the correlation between mask-wearing and other protective behaviours [1, 15]. We investigate this phenomena by assessing how much effectiveness estimates change when previously observed factors are excluded, following Sharma *et al*. [18].

Figures 12 and 13 show NPI effectiveness estimates when each observed NPI is excluded in turn. Figure 14 shows the sensitivity of our effect estimates to excluding mobility from our model. Reducing mobility has a large effect on *R*, so it is encouraging to see that our effects are robust to excluding mobility from our model.

**Fig. 12.**
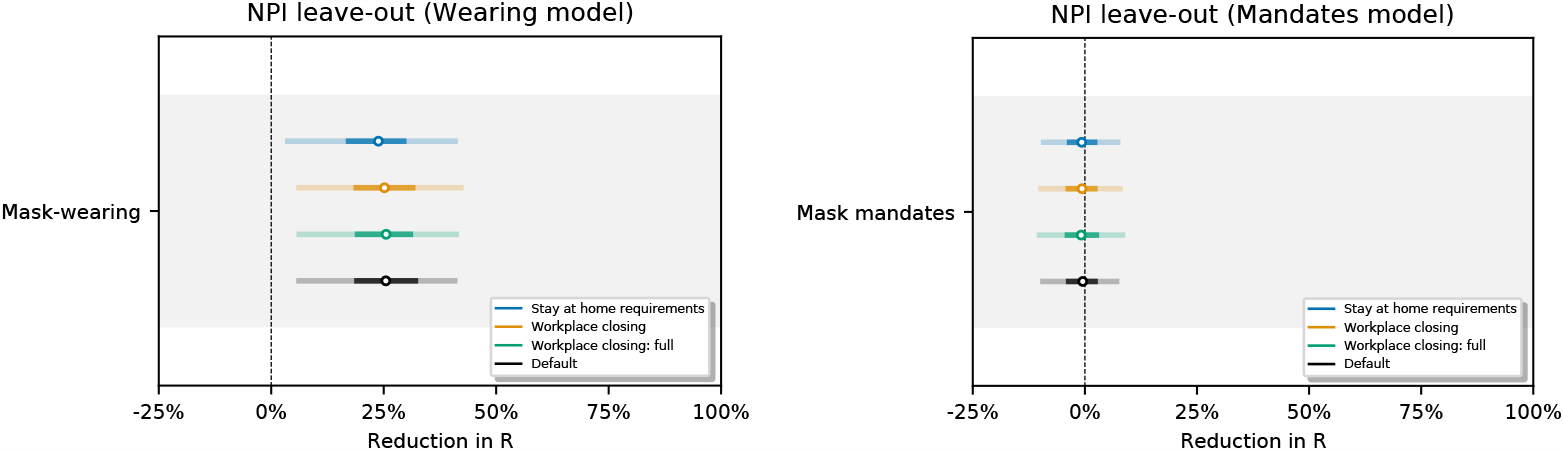
Sensitivity of our effect estimates to leaving out recorded interventions, simulating unobserved confounding effects on transmission.

**Fig. 13.**
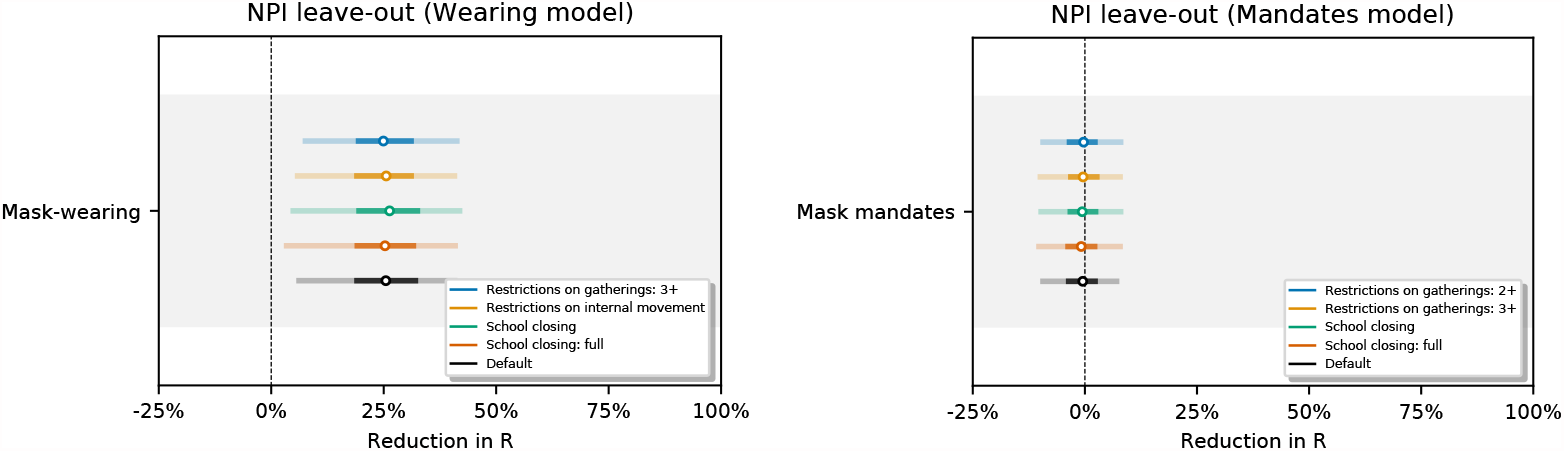
Round 2 (different NPIs left out): Sensitivity of our effect estimates to leaving out recorded interventions, simulating unobserved confounding effects on transmission.

**Fig. 14.**
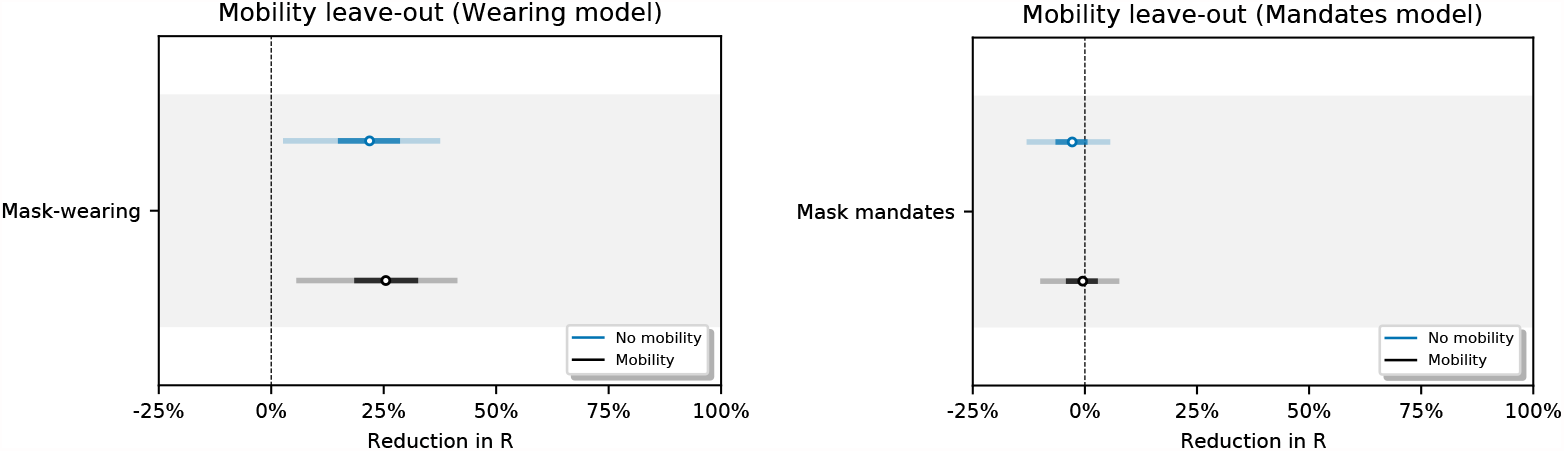
Sensitivity of effect estimates to excluding mobility from our model.

One objection to our methodology is that mask-wearing increases over our window of analysis while transmission decreases in many regions. It is therefore possible that this correlation is a spurious contributor to the substantial apparent wearing effect. We test this hypothesis by creating a fake wearing variable for each region. Each variable has the same start and end wearing value as the true wearing percentage and linearly interpolates between these values to capture the trend in wearing in that region. We infer a small and uncertain effect for the fake wearing variable 7.6% [–20.2%, 30.0%] (see Figure 15). This implies that the wearing effect we infer does not rely solely on the wearing trend in this period. Figure 16 shows the sensitivity of our effect estimates to excluding all NPIs from our wearing model.

**Fig. 15.**
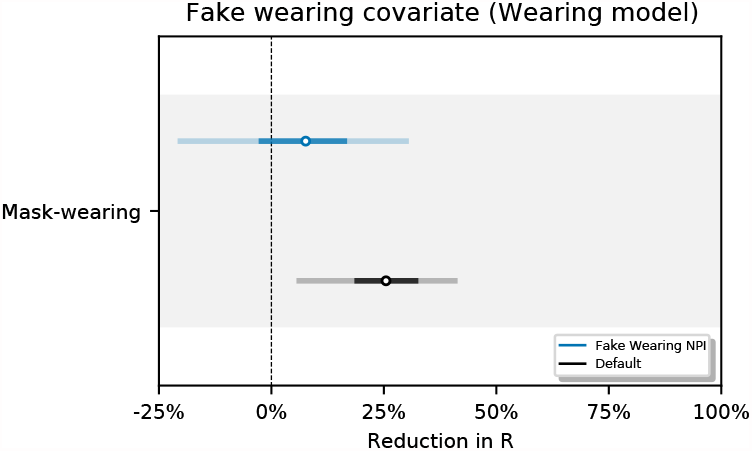
Effect estimates when wearing data is replaced by synthetic data that tracks the linear change in wearing, in our window, for each region.

**Fig. 16.**
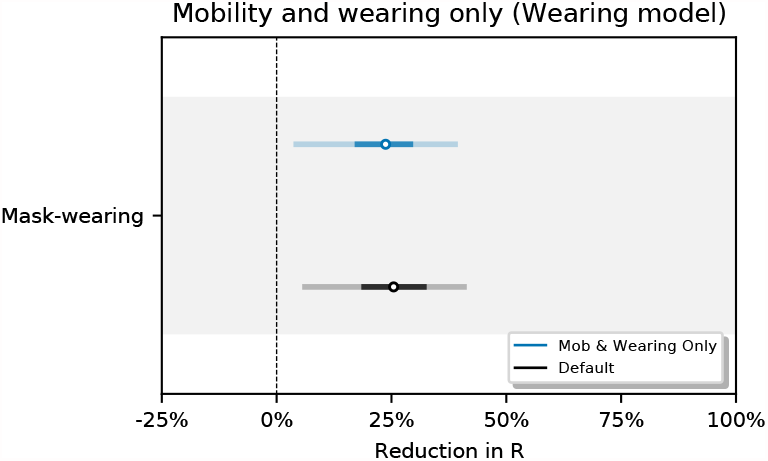
Sensitivity of effect estimates to excluding all NPIs.

#### C.2 Epidemiological priors

Figure 17 shows the sensitivity of our effect estimates to 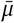, the mean of the prior over *μ* in 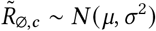, where *μ* ∼ TruncatedNormal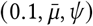. Recall that 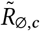 is the reproduction number at the start of the window of analysis, supposing mandates are not active and no one is wearing masks. Figure 18 shows the sensitivity of our effect estimates to *ψ* the scale of the prior over *μ* in 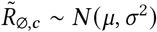, where *μ* ∼ TruncatedNormal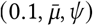. Figure 19 shows the sensitivity of our effect estimates to *ω*, the scale of the prior over *σ* in 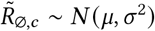, where *σ* ∼ HalfNormal (*ω*). Figure 20 shows the sensitivity of our effect estimates to the prior over the random walk noise scale.

**Fig. 17.**
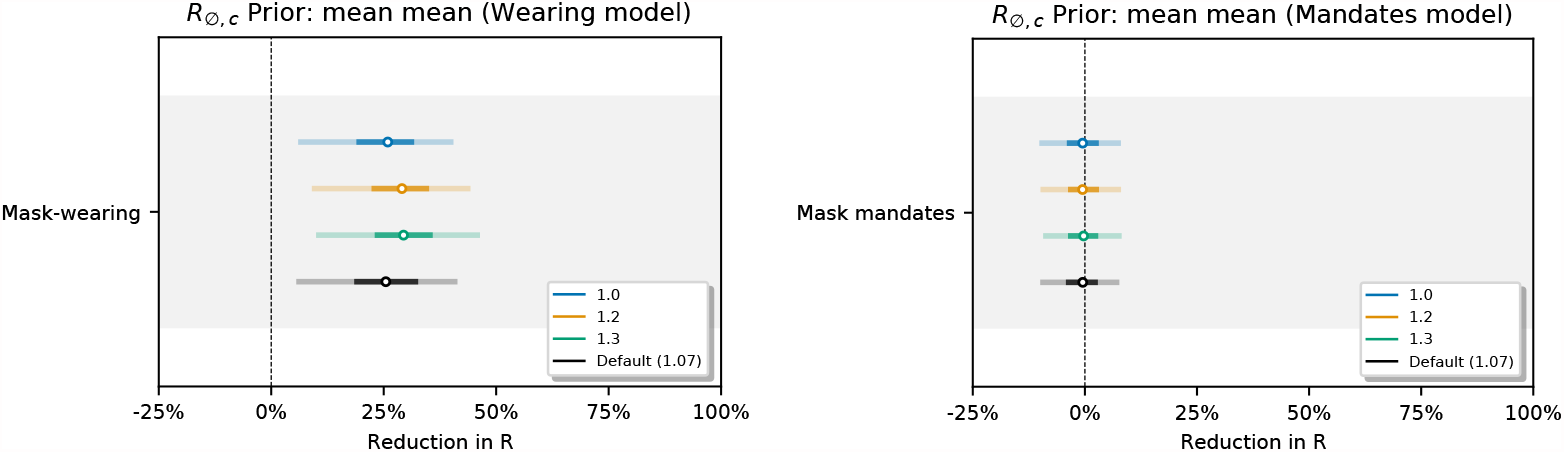
Sensitivity of effect estimates to 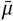, the mean of the prior over *μ* in 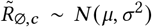, where *σ* ∼ TruncatedNormal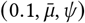. (L): wearing, (R): mandates.

**Fig. 18.**
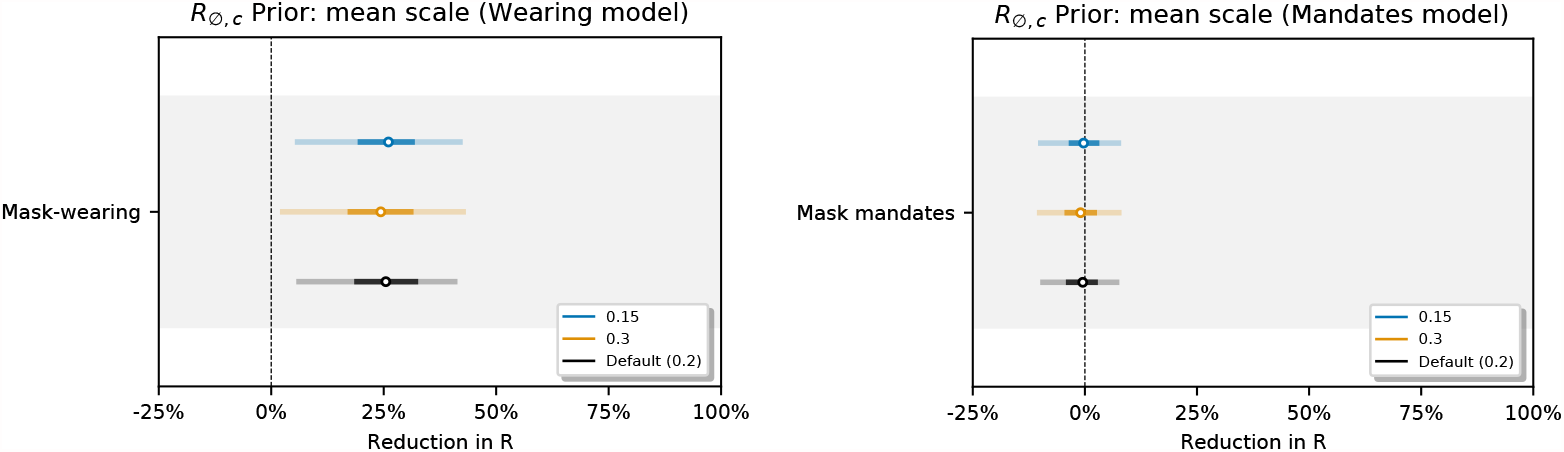
Sensitivity of our effect estimates to *ψ* the scale of the prior over *μ* in 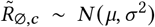, where *σ* ∼ TruncatedNormal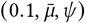.

**Fig. 19.**
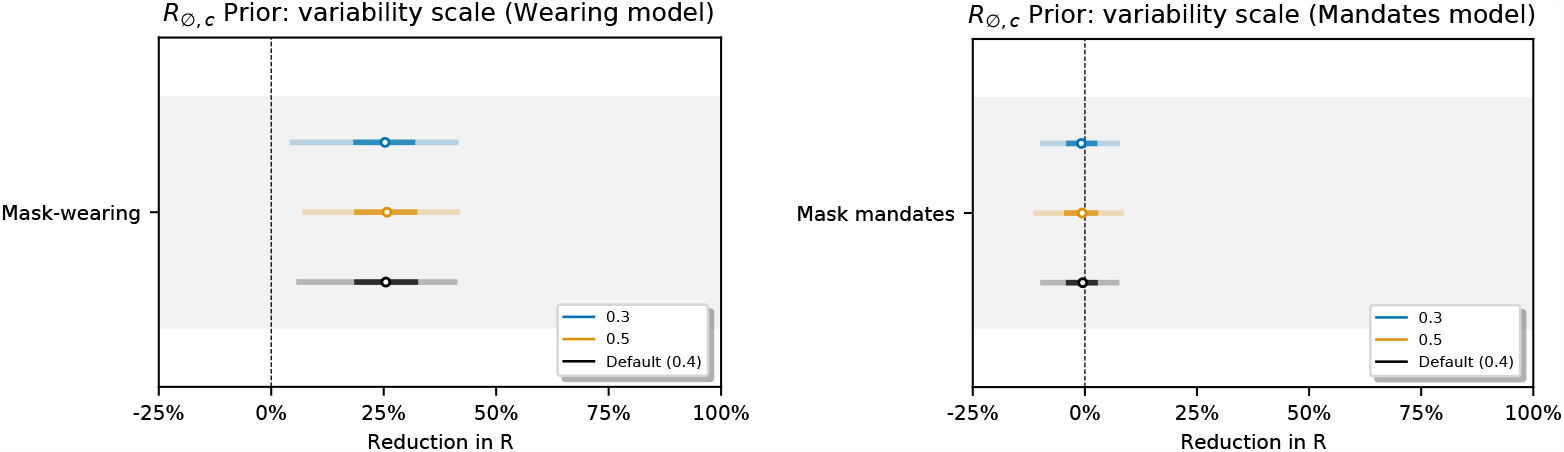
Sensitivity of our effect estimates to *ω*, the scale of the prior over *σ* in 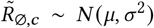, where *σ* ∼ HalfNormal(*ω*).

**Fig. 20.**
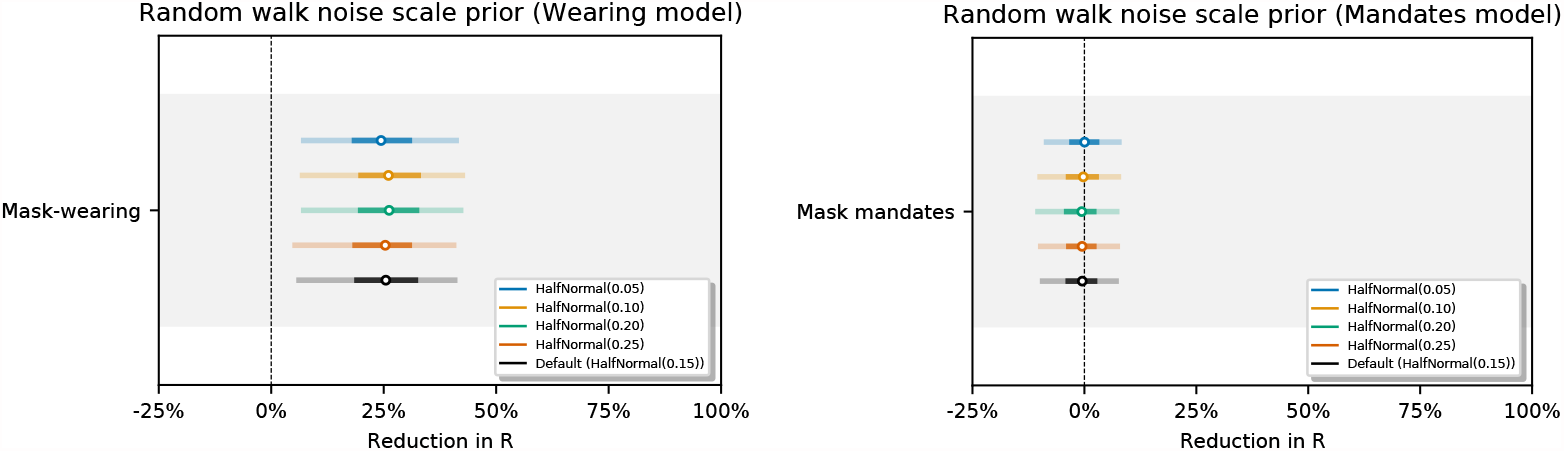
Sensitivity of our effect estimates to the noise scale of the weekly random walk.

#### C.3 Delay distributions

Figure 21 shows the sensitivity of the effect estimates to the mean of the distribution of the generation interval. Figures 22 and 23 show the sensitivity of the effect estimates to the mean and dispersion of the distribution that represents the delay between infection and case reporting.

**Fig. 21.**
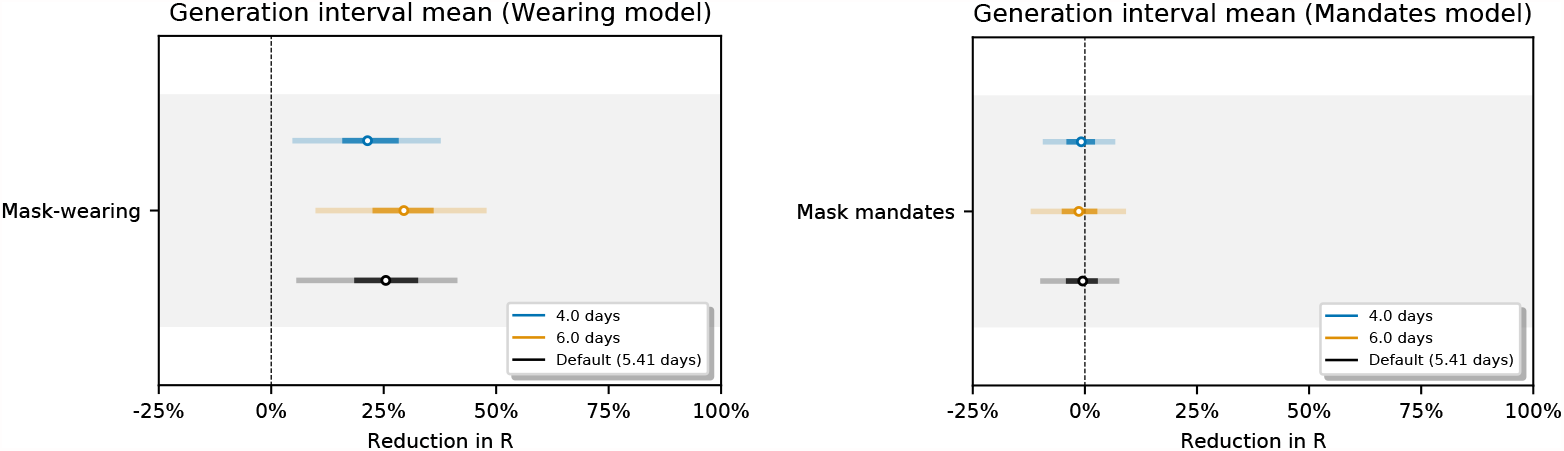
Sensitivity of our effect estimates to the mean of the generation interval.

**Fig. 22.**
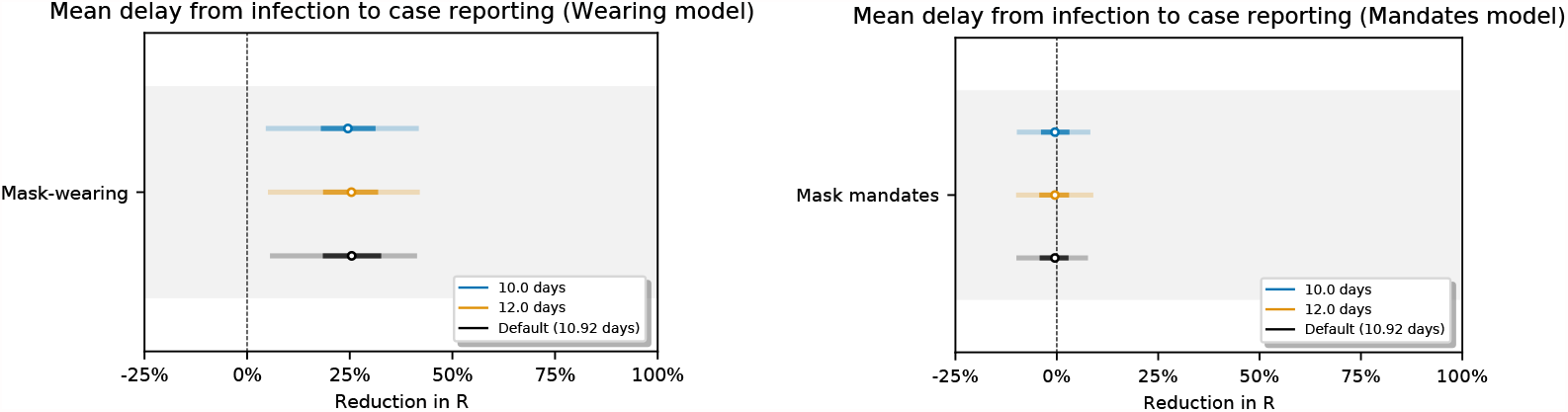
Sensitivity of our effect estimates to the mean of the delay from infection to case reporting.

**Fig. 23.**
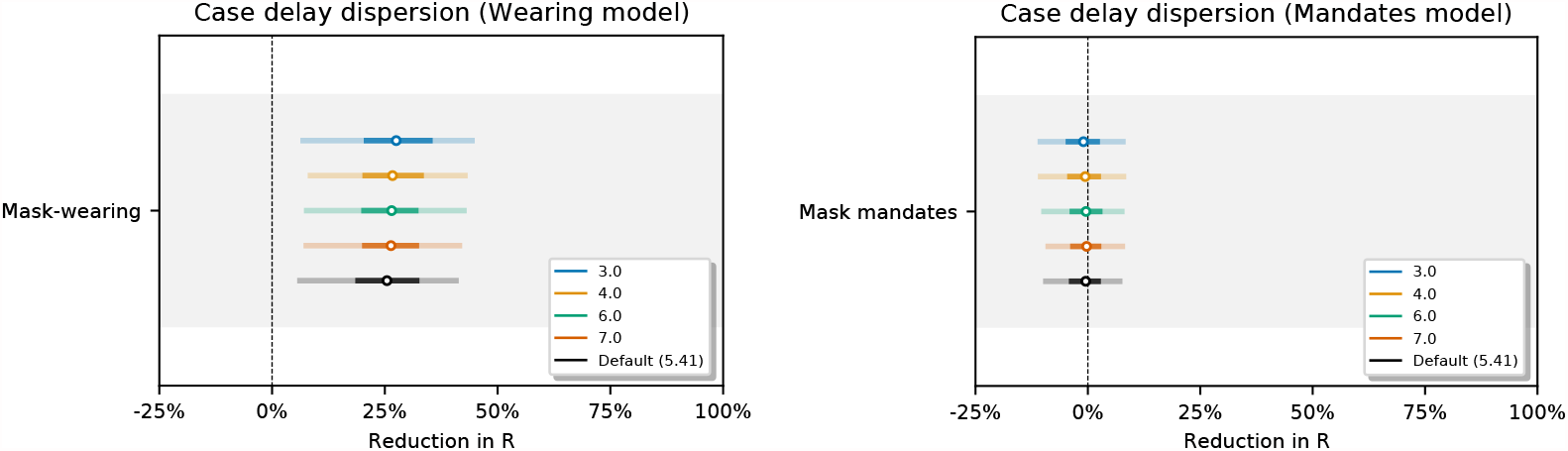
Sensitivity of our effect estimates to the dispersion of the delay from infection to case reporting.

#### C.4 Covariate priors

Figure 24 shows the sensitivity of our effect estimates to the prior over the NPI effects. Figure 25 shows the sensitivity of our effect estimates to the scale of the prior over the wearing effect. Figure 26 shows the sensitivity of our effect estimates to the scale of the prior over the mandate effect. Figure 27 shows the sensitivity of our effect estimates to the mean of the prior over the mobility effect.

**Fig. 24.**
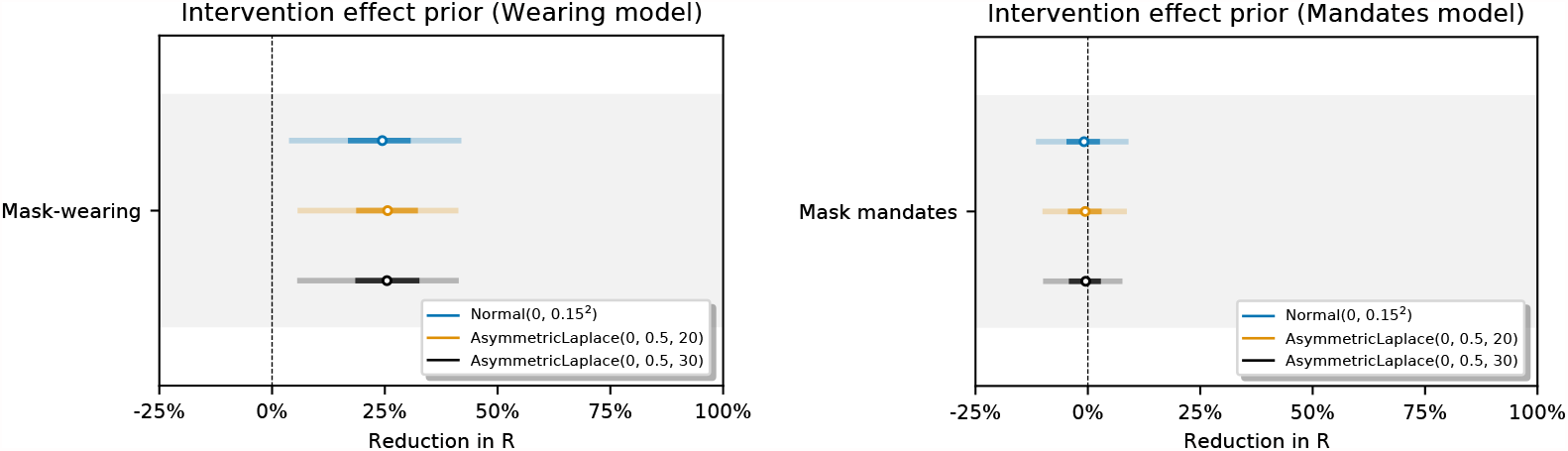
Sensitivity of our effect estimates to the prior over the NPI effects.

**Fig. 25.**
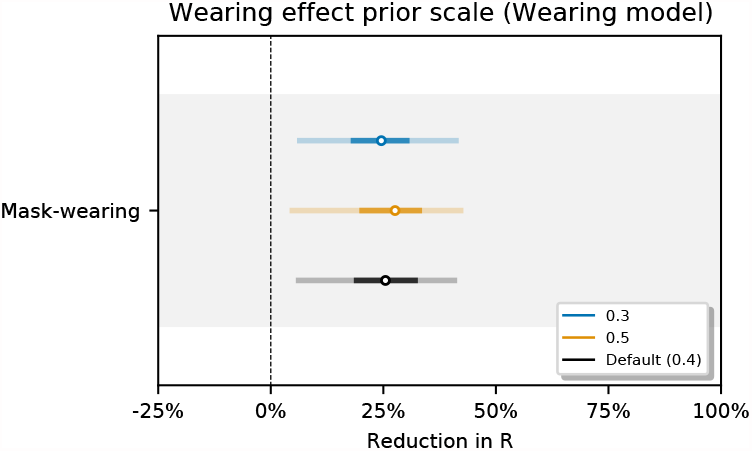
Sensitivity of our effect estimates to the scale of the prior over the wearing effect.

**Fig. 26.**
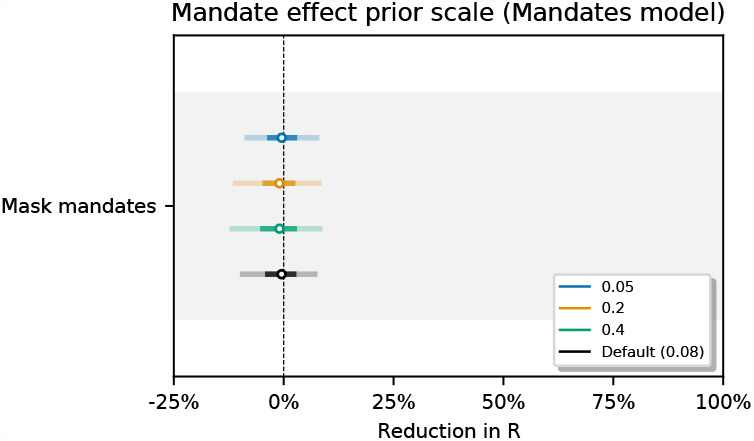
Sensitivity of our effect estimates to the scale of the prior over the mandate effect.

**Fig. 27.**
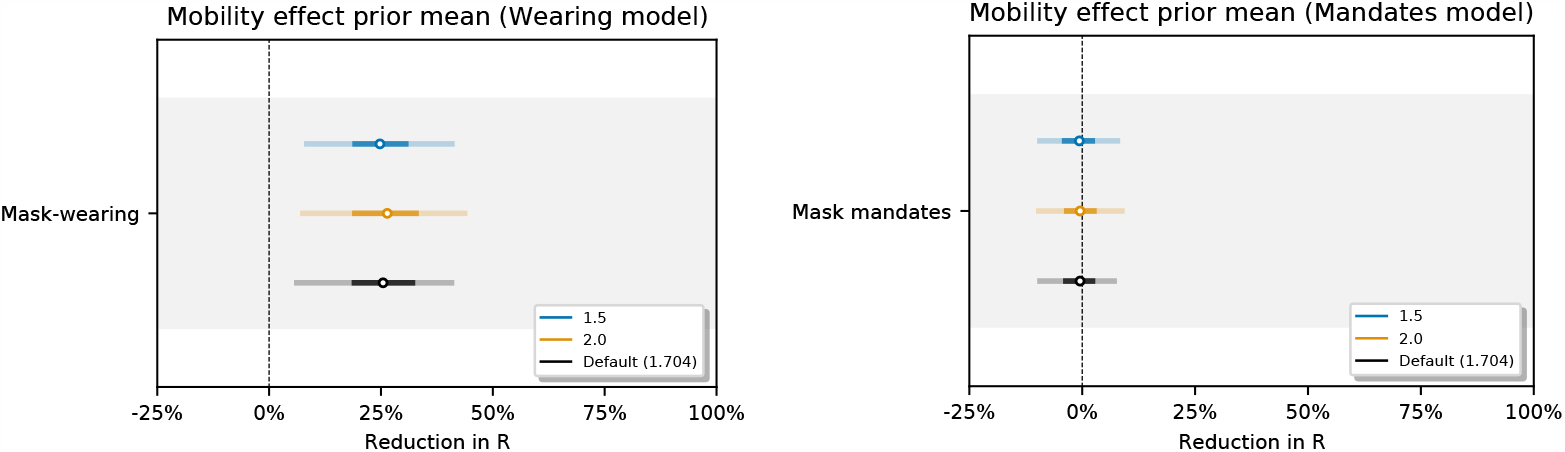
Sensitivity of our effect estimates to the mean of the prior over the mobility effect.

Figure 28 shows the sensitivity of our effect estimates to the scale of the prior over the mobility effect.

**Fig. 28.**
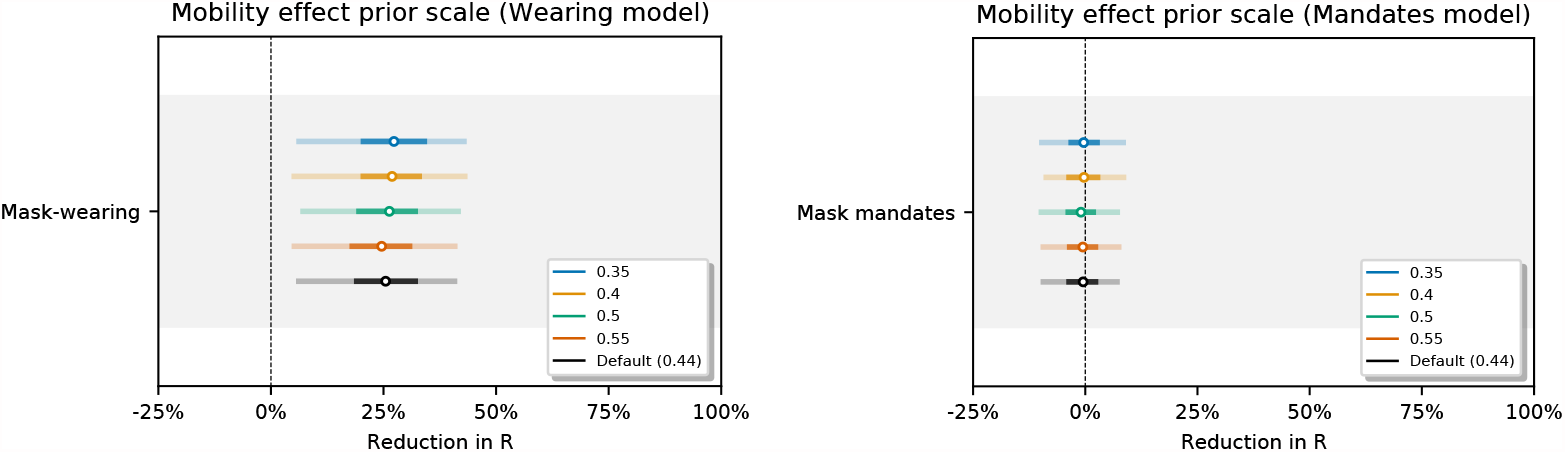
Sensitivity of our effect estimates to the scale of the prior over the mobility effect.

#### C.5 Model structure

Figure 29 shows the sensitivity of our effect estimates to the parameterisation of the wearing effect. The wearing parameterisations are defined as follows:

**Fig. 29.**
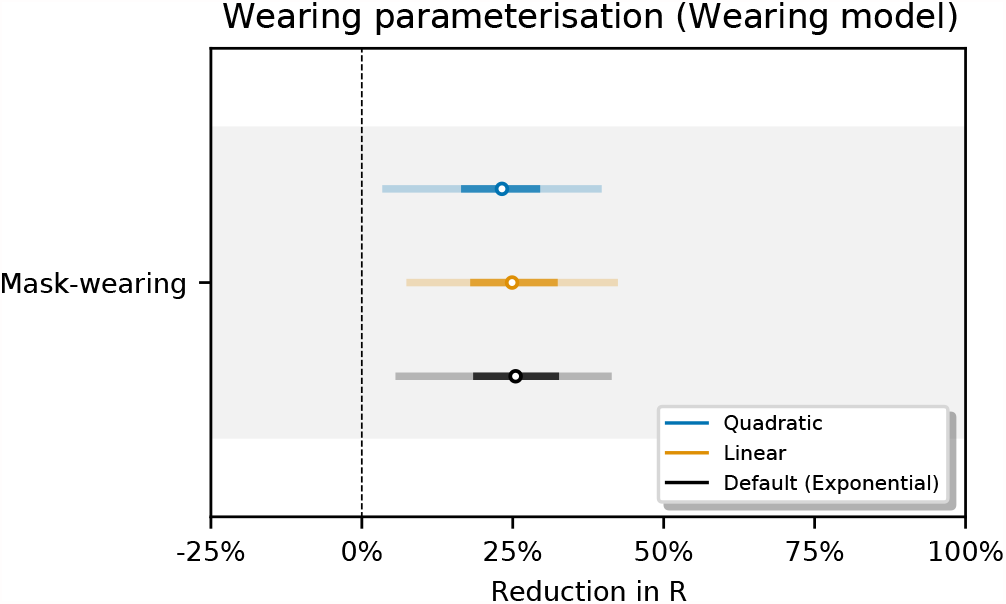
Sensitivity of our effect estimates to the to the parameterisation of the wearing effect.

- *Exponential* (base model): 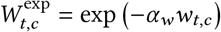. We use this form in our base model because it is consistent with the form of the mandate effect on *R*.
- *Linear*: 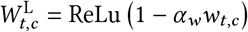, where ReLU is the Rectified Linear Unit. The ReLU function preserves positive inputs and maps negative inputs to zero. We include the linear form because it is the simplest way to approximate wearing’s effect on transmission.
- *Quadratic*: 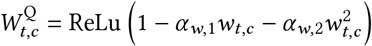. The quadratic form is based on a simple model: suppose two people interact, and there is a fixed, independent probability that each of them wears a mask. Then the reduction in the probability of transmission due to mask-wearing is quadratic in the probability that each wears a mask. The two *α* parameters correspond to source control and wearer-protection.

Figure 30 shows the sensitivity of our effect estimates to the period of the random walk. For a period of N days the value of *R*_*t,c*_ may change without a change of covariates every N days.

**Fig. 30.**
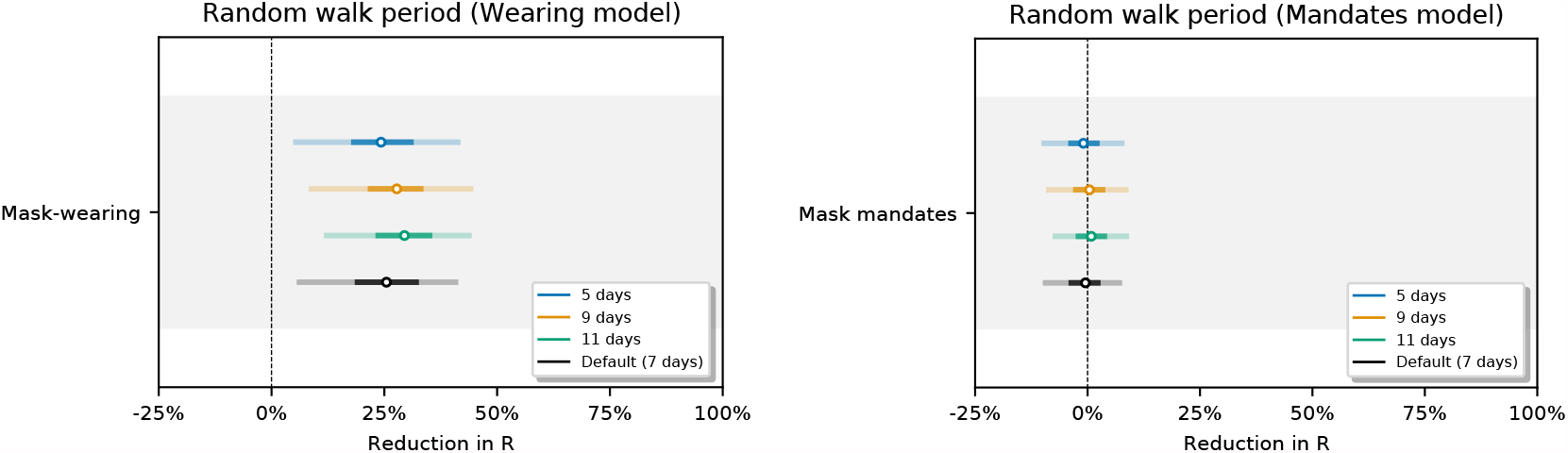
Sensitivity of our effect estimates to the period of the random walk.

#### C.6 Data permutations

Figures 31, 32 and 33 show the sensitivity of our effect estimates to bootstrapping our regions. Bootstrapping assesses how much our effect estimates depend on the regions we included. For each seed we sample 92 regions with replacement from our set of 92 regions. Each bootstrap contains 58/92 *unique* regions on average.

**Fig. 31.**
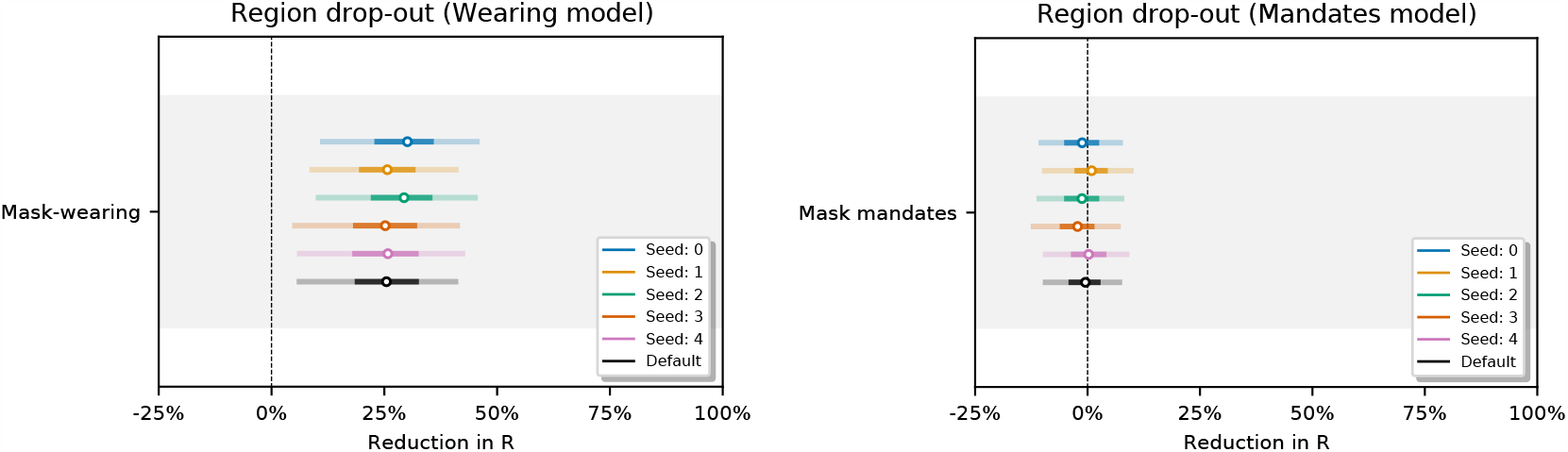
Sensitivity of our effect estimates using random bootstrapped sets of regions. Seed 0-4.

**Fig. 32.**
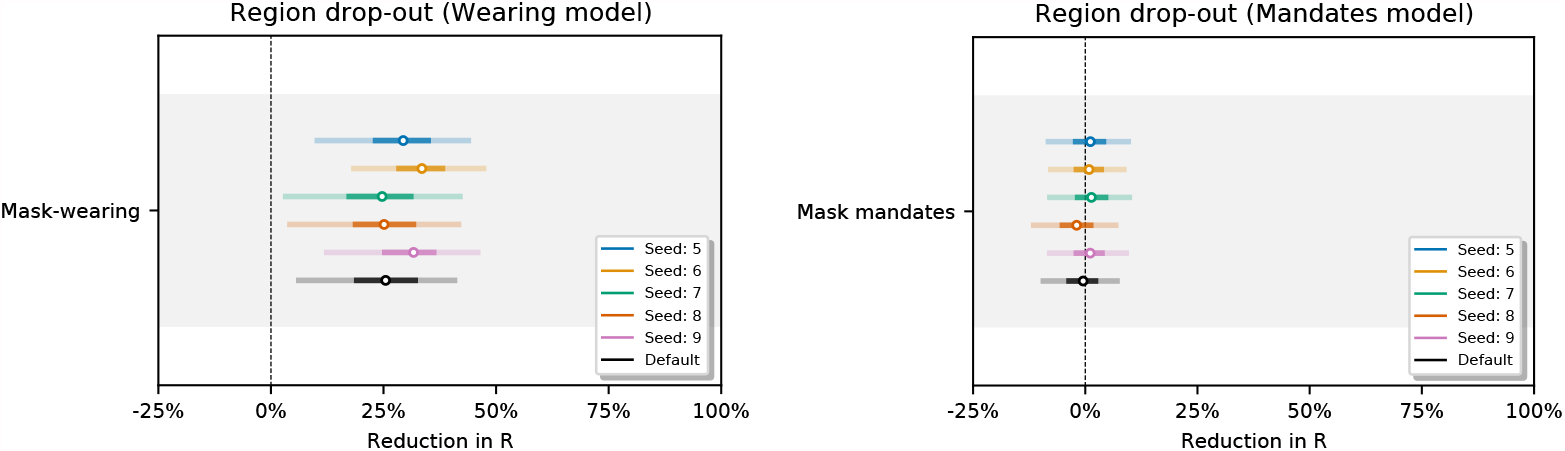
Sensitivity of our effect estimates using random bootstrapped sets of regions. Seed 5-9.

**Fig. 33.**
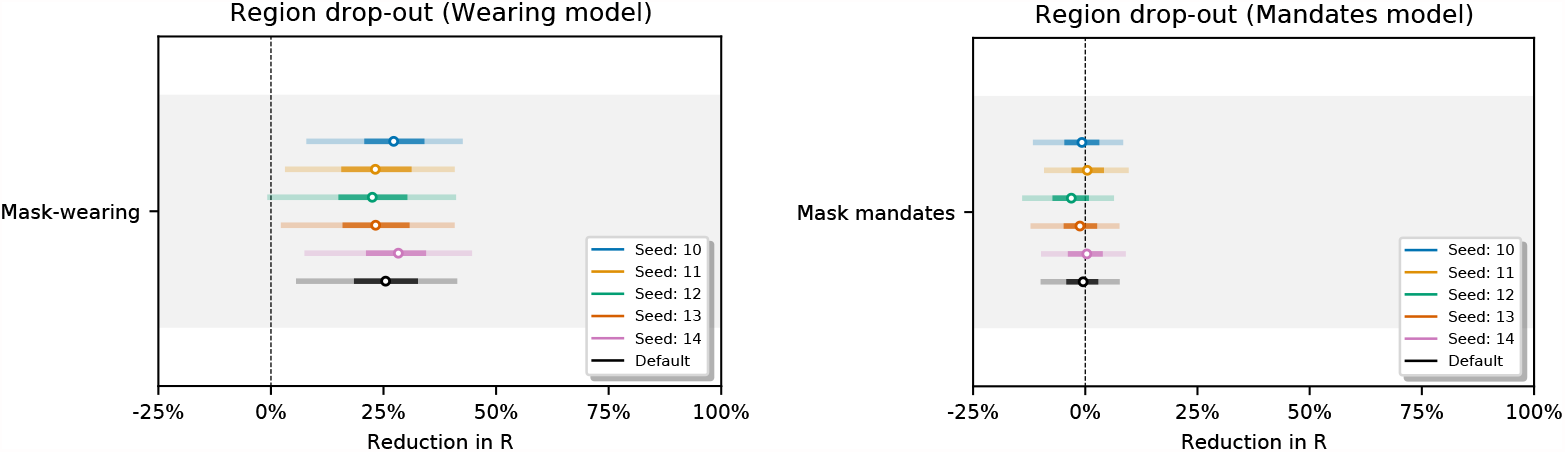
Sensitivity of our effect estimates using random bootstrapped sets of regions. Seed 10-14.

Figure 34 shows the sensitivity of our effect estimates to assuming a persistent mandate effect that lasts beyond the point the mandate is lifted. Figure 35 shows the sensitivity of our effect estimates when removing the less stringent mask mandate feature. Figure 36 shows the sensitivity of our effect estimates to shorter periods of analysis. We see little variation in our effect estimates, which implies that our results may generalise to other periods.

**Fig. 34.**
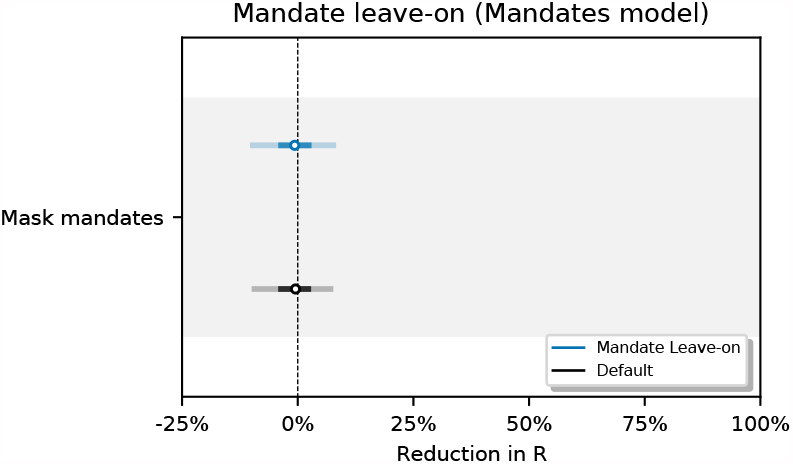
Sensitivity of our effect estimates to assuming a persistent mandate effect that lasts beyond the point the mandate is lifted.

**Fig. 35.**
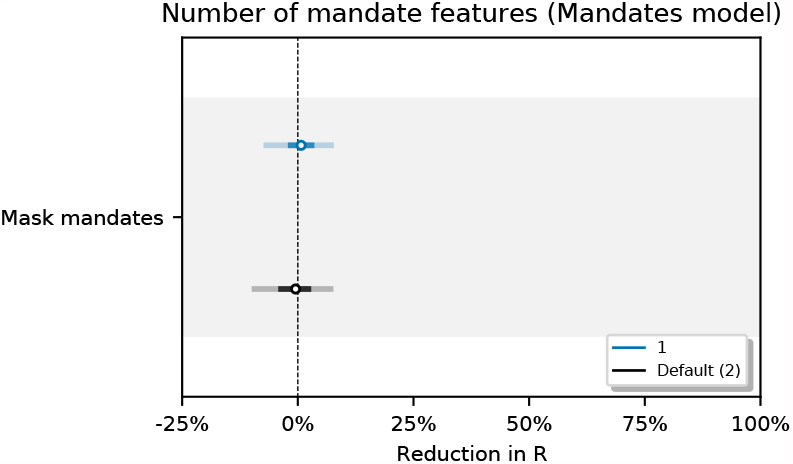
Sensitivity of our effect estimates when removing the less stringent mask mandate feature.

**Fig. 36.**
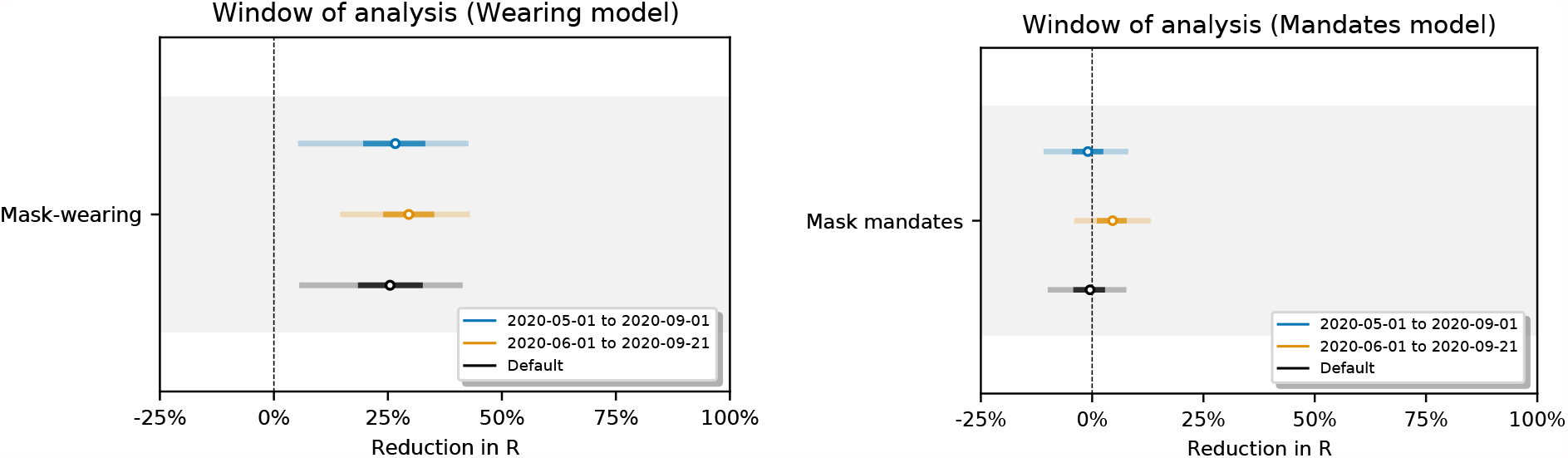
Sensitivity of our effect estimates to the window of analysis.

## Footnotes

1 All confidence or credible intervals reported in this text are 95% intervals unless stated otherwise.

2 A hyperprior is a prior distribution placed a parameter describing another prior distribution.

